# Duration of mRNA vaccine protection against SARS-CoV-2 Omicron BA.1 and BA.2 subvariants in Qatar

**DOI:** 10.1101/2022.03.13.22272308

**Authors:** Hiam Chemaitelly, Houssein H. Ayoub, Sawsan AlMukdad, Peter Coyle, Patrick Tang, Hadi M. Yassine, Hebah A. Al-Khatib, Maria K. Smatti, Mohammad R. Hasan, Zaina Al-Kanaani, Einas Al- Kuwari, Andrew Jeremijenko, Anvar Hassan Kaleeckal, Ali Nizar Latif, Riyazuddin Mohammad Shaik, Hanan F. Abdul-Rahim, Gheyath K. Nasrallah, Mohamed Ghaith Al-Kuwari, Adeel A. Butt, Hamad Eid Al-Romaihi, Mohamed H. Al-Thani, Abdullatif Al-Khal, Roberto Bertollini, Laith J. Abu-Raddad

## Abstract

The SARS-CoV-2 Omicron (B.1.1.529) variant has two subvariants, BA.1 and BA.2, that are genetically quite divergent. We conducted a matched, test-negative, case-control study to estimate duration of protection of mRNA COVID-19 vaccines, after the second dose and after a third/booster dose, against BA.1 and BA.2 infections in Qatar’s population. BNT162b2 effectiveness against symptomatic BA.1 infection was highest at 46.6% (95% CI: 33.4-57.2%) in the first three months after the second dose, but then declined to ∼10% or below thereafter. Effectiveness rapidly rebounded to 59.9% (95% CI: 51.2-67.0%) in the first month after the booster dose, but then started to decline again. BNT162b2 effectiveness against symptomatic BA.2 infection was highest at 51.7% (95% CI: 43.2-58.9%) in the first three months after the second dose, but then declined to ∼10% or below thereafter. Effectiveness rapidly rebounded to 43.7% (95% CI: 36.5-50.0%) in the first month after the booster dose, but then declined again. Effectiveness against COVID-19 hospitalization and death was in the range of 70-80% any time after the second dose, and was greater than 90% after the booster dose. Similar patterns of protection were observed for the mRNA-1273 vaccine. mRNA vaccines provide only moderate and short-lived protection against symptomatic Omicron infections, with no discernable differences in protection against either the BA.1 or BA.2 subvariants. Vaccine protection against COVID-19 hospitalization and death is strong and durable after the second dose, but is more robust after a booster dose.

## Introduction

Qatar endured a severe acute respiratory syndrome coronavirus 2 (SARS-CoV-2) Omicron (B.1.1.529)^1^ wave that started on December 19, 2021 and peaked in mid-January, 2022^2-5^. The wave was first dominated by the BA.1 Omicron subvariant, but within a few days, the BA.2 subvariant predominated (Figure 1). While BA.1 and BA.2 remain classified as subvariants of the Omicron variant, there is considerable genetic distance between them^6^. Accordingly, we investigated duration of protection of BNT162b2 (Pfizer-BioNTech)^7^ and mRNA-1273 (Moderna)^8^ mRNA coronavirus disease 2019 (COVID-19) vaccines, after the second dose and after the third/booster dose, against symptomatic BA.1 and BA.2 infections, between December 23, 2021 and February 28, 2022. Duration of vaccine protection was also investigated against any severe (acute-care hospitalization)^9^, critical (intensive-care-unit hospitalization)^9^, or fatal^10^ infection due to either Omicron subvariant.

**Figure 1.**
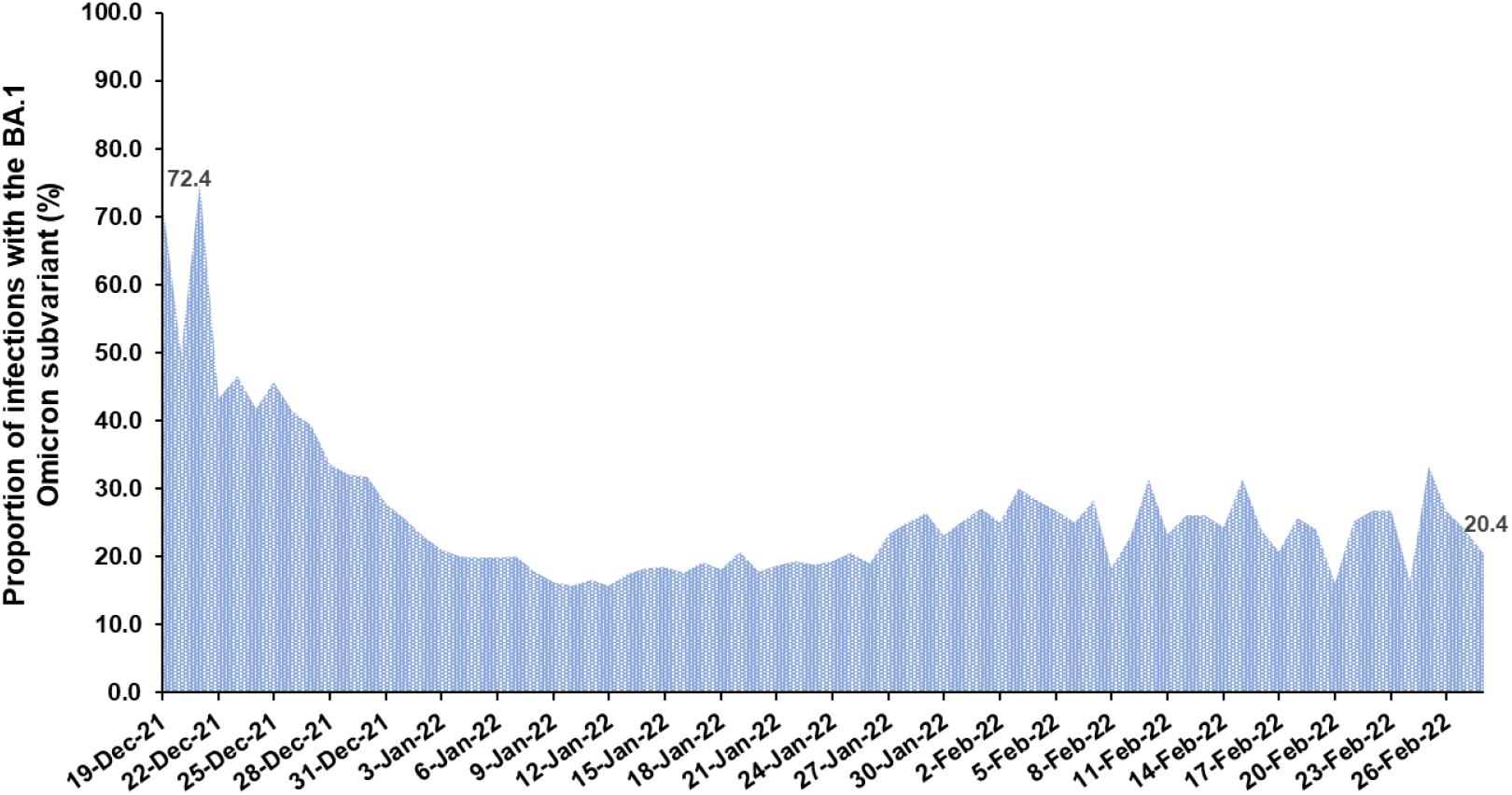
Proportion of Omicron infections with the BA.1 (versus BA.2) subvariant in PCR-positive tests assessed using TaqPath COVID-19 Combo Kit during the study period.

Vaccine effectiveness was estimated using the test-negative, case-control study design^11,12^, applying methodology that was developed earlier to assess duration of protection of the BNT162b2^13^ and mRNA-1273^14^ vaccines in the same population during pre-Omicron SARS-CoV-2 infection waves (Methods). Cases (persons infected with BA.1, BA.2, or any-Omicron-subvariant) and controls (uninfected persons) were exact-matched by sex, 10-year age group, nationality, and calendar week of polymerase chain reaction (PCR) test to control for established differences in the risk of exposure to SARS-CoV-2 infection in Qatar^15-19^.

## Results

### Main analyses

By February 28, 2022 (end of study), 1,308,926 individuals received 2 or more BNT162b2 doses, and 355,979 of these received a booster dose. Meanwhile, 894,142 individuals received 2 or more mRNA-1273 doses, and 146,961 of these received a booster dose. The median dates of first, second, and third doses were May 3, 2021, May 24, 2021, and December 27, 2021 for BNT162b2; and May 28, 2021, June 27, 2021, and January 16, 2022 for mRNA-1273, respectively. The median time between the first and second doses was 21 days (interquartile range (IQR), 21-22 days) for BNT162b2 and 28 days (IQR, 28-30 days) for mRNA-1273. The median time between the second and booster doses was 251 days (IQR, 233-275 days) for BNT162b2 and 236 days (IQR, 213-261 days) for mRNA-1273.

The process used to select the study populations is shown in Figure 2. Demographic characteristics of the study populations are presented in Tables 1 and 2. The study was conducted based on the total population of Qatar. The study populations are therefore representative of the internationally diverse, but predominantly young and male population of Qatar.

**Figure 2.**
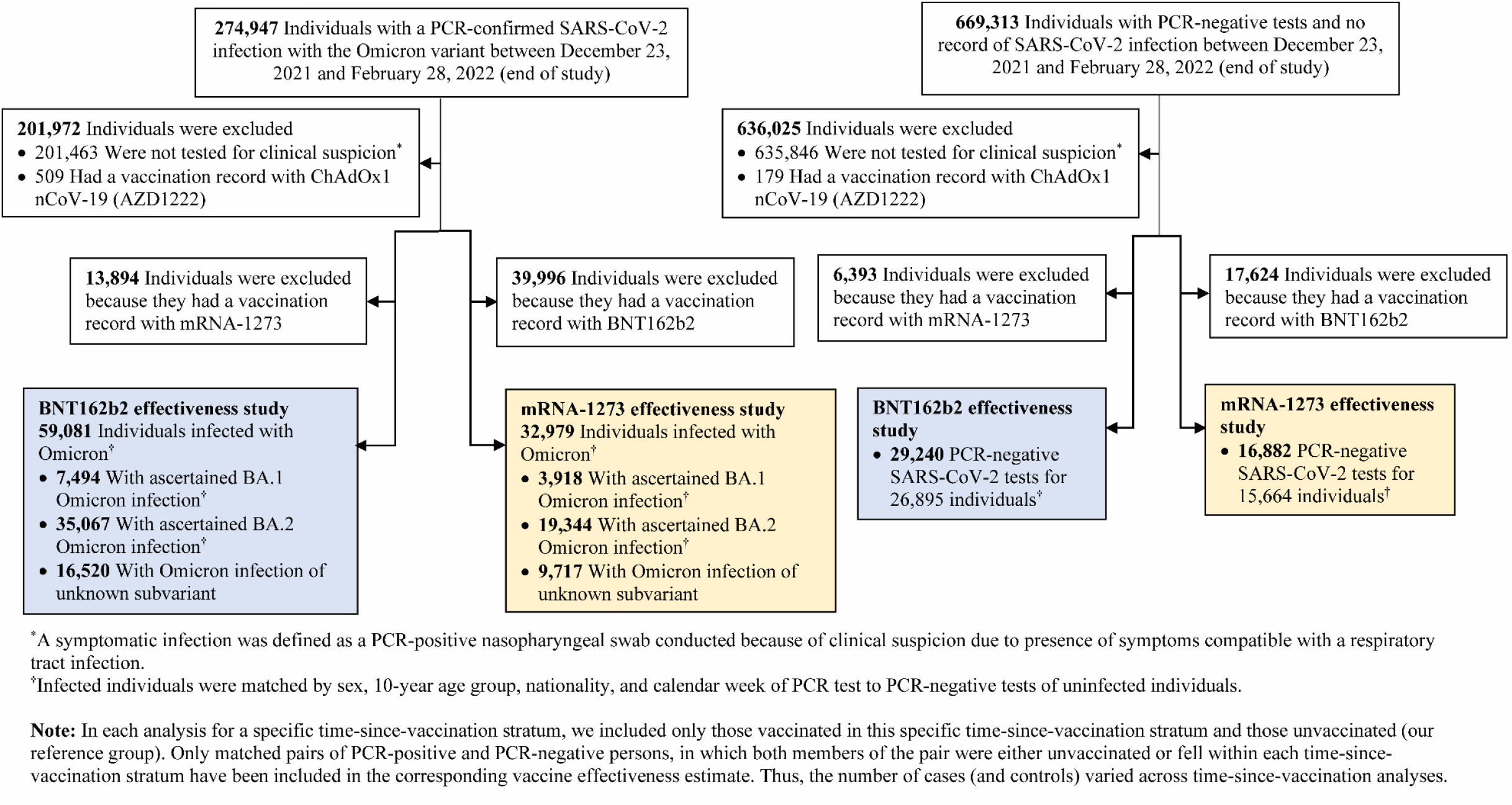
Flowchart describing the population selection process for investigating effectiveness of the BNT162b2 and mRNA-1273 vaccines during the SARS-CoV-2 Omicron infection wave.

**Table 1.** Demographic characteristics of cases and controls in samples used to estimate effectiveness of the BNT162b2 vaccine against symptomatic SARS-CoV-2 BA.1 Omicron infection, symptomatic BA.2 Omicron infection, and any symptomatic Omicron infection. The table was generated by combining the matched samples of the various time-since-vaccination strata.

| Characteristics | Effectiveness against symptomatic SARS-CoV-2<br>BA.1 Omicron infection |  |  | Effectiveness against symptomatic SARS-CoV-2<br>BA.2 Omicron infection |  |  | Effectiveness against any symptomatic SARS-CoV-2 Omicron infection |  |  |
| --- | --- | --- | --- | --- | --- | --- | --- | --- | --- |
|  | Cases*<br>(PCR-positive) | Controls*<br>(PCR-negative) | SMD† | Cases‡<br>(PCR-positive) | Controls‡<br>(PCR-negative) | SMD† | Cases§<br>(PCR-positive) | Controls§<br>(PCR-negative) | SMD† |
|  | N=7,072 | N=12,278 |  | N=21,541 | N=21,541 |  | N=39,855 | N=23,814 |  |
| **Median age (IQR) — years** | 32 (22-42) | 32 (20-42) | 0.06† | 32 (18-42) | 31 (18-42) | 0.01† | 32 (19-42) | 32 (19-43) | 0.01† |
| **Age group — no. (%)** |  |  |  |  |  |  |  |  |  |
| <10 years | 670 (9.5) | 1,282 (10.4) | 0.05 | 3,157 (14.7) | 3,157 (14.7) | 0.00 | 5,581 (14.0) | 3,501 (14.7) | 0.03 |
| 10-19 years | 875 (12.5) | 1,611 (13.1) |  | 2,458 (11.4) | 2,458 (11.4) |  | 4,594 (11.5) | 2,640 (11.1) |  |
| 20-29 years | 1,385 (19.7) | 2,389 (19.5) |  | 4,016 (18.6) | 4,016 (18.6) |  | 7,344 (18.4) | 4,369 (18.4) |  |
| 30-39 years | 1,983 (28.2) | 3,487 (28.4) |  | 5,561 (25.8) | 5,561 (25.8) |  | 10,419 (26.1) | 6,066 (25.5) |  |
| 40-49 years | 1,053 (15.0) | 1,782 (14.5) |  | 2,824 (13.1) | 2,824 (13.1) |  | 5,462 (13.7) | 3,254 (13.7) |  |
| 50-59 years | 674 (9.6) | 1,115 (9.1) |  | 2,166 (10.1) | 2,166 (10.1) |  | 3,995 (10.0) | 2,440 (10.3) |  |
| 60-69 years | 279 (4.0) | 436 (3.6) |  | 951 (4.4) | 951 (4.4) |  | 1,685 (4.2) | 1,050 (4.4) |  |
| 70+ years | 103 (1.5) | 176 (1.4) |  | 408 (1.9) | 408 (1.9) |  | 775 (1.9) | 494 (2.1) |  |
| **Sex** |  |  |  |  |  |  |  |  |  |
| Male | 3,437 (49.0) | 6,335 (51.6) | 0.05 | 11,986 (55.6) | 11,986 (55.6) | 0.00 | 21,951 (55.1) | 13,257 (55.7) | 0.01 |
| Female | 3,585 (51.1) | 5,943 (48.4) |  | 9,555 (44.4) | 9,555 (44.4) |  | 17,904 (44.9) | 10,557 (44.3) |  |
| **Nationality\*\*** |  |  |  |  |  |  |  |  |  |
| Bangladeshi | 102 (1.5) | 184 (1.5) | 0.05 | 521 (2.4) | 521 (2.4) | 0.00 | 872 (2.2) | 614 (2.6) | 0.06 |
| Egyptian | 416 (5.9) | 723 (5.9) |  | 1,384 (6.4) | 1,384 (6.4) |  | 2,360 (5.9) | 1,343 (5.6) |  |
| Filipino | 761 (10.8) | 1,357 (11.1) |  | 2,063 (9.6) | 2,063 (9.6) |  | 3,844 (9.6) | 2,227 (9.4) |  |
| Indian | 793 (11.3) | 1,467 (12.0) |  | 3,077 (14.3) | 3,077 (14.3) |  | 5,403 (13.6) | 3,314 (13.9) |  |
| Nepalese | 80 (1.1) | 138 (1.1) |  | 430 (2.0) | 430 (2.0) |  | 632 (1.6) | 369 (1.6) |  |
| Pakistani | 152 (2.2) | 279 (2.3) |  | 788 (3.7) | 788 (3.7) |  | 1,325 (3.3) | 805 (3.4) |  |
| Qatari | 2,824 (40.2) | 5,074 (41.3) |  | 7,277 (33.8) | 7,277 (33.8) |  | 14,632 (36.7) | 8,304 (34.9) |  |
| Sri Lankan | 62 (0.9) | 105 (0.9) |  | 299 (1.4) | 299 (1.4) |  | 497 (1.3) | 313 (1.3) |  |
| Sudanese | 328 (4.7) | 576 (4.7) |  | 1,036 (4.8) | 1,036 (4.8) |  | 1,730 (4.3) | 1,026 (4.3) |  |
| Other nationalities | 1,504 (21.4) | 2,375 (19.3) |  | 4,666 (21.7) | 4,666 (21.7) |  | 8,560 (21.5) | 5,499 (23.1) |  |

Abbreviations: IQR, interquartile range; PCR, polymerase chain reaction; SMD, standardized mean difference.
†Cases and controls were matched one-to-two by sex, 10-year age group, nationality, and calendar week of PCR test.
‡SMD is the difference in the mean of a covariate between groups divided by the pooled standard deviation. An SMD<0.1 indicates adequate matching.
§Cases and controls were matched one-to-one by sex, 10-year age group, nationality, and calendar week of PCR test.
¶Cases and controls were matched two-to-one by sex, 10-year age group, nationality, and calendar week of PCR test.
\*\*SMD is for the mean difference between groups divided by the pooled standard deviation.
\*\*\*Nationalities were chosen to represent the most populous groups in Qatar.

**Table 2.** Demographic characteristics of cases and controls in samples used to estimate effectiveness of the mRNA-1273 vaccine against symptomatic SARS-CoV-2 BA.1 Omicron infection, symptomatic BA.2 Omicron infection, and any symptomatic Omicron infection. The table was generated by combining the matched samples of the various time-since-vaccination strata.

| Characteristics | Effectiveness against symptomatic SARS-CoV-2<br>BA.1 Omicron infection |  |  | Effectiveness against symptomatic SARS-CoV-2<br>BA.2 Omicron infection |  |  | Effectiveness against any symptomatic SARS-CoV-2 Omicron infection |  |  |
| --- | --- | --- | --- | --- | --- | --- | --- | --- | --- |
|  | Cases*<br>(PCR-positive) | Controls*<br>(PCR-negative) | SMD† | Cases*<br>(PCR-positive) | Controls*<br>(PCR-negative) | SMD† | Cases*<br>(PCR-positive) | Controls*<br>(PCR-negative) | SMD† |
|  | N=3,574 | N=6,176 |  | N=13,537 | N=13,537 |  | N=21,810 | N=13,288 |  |
| **Median age (IQR) — years** | 30 (15-38) | 29 (11-38) | 0.07‡ | 28 (10-37) | 28 (10-38) | 0.01‡ | 28 (9-37) | 28 (9-38) | 0.00‡ |
| **Age group — no. (%)** |  |  |  |  |  |  |  |  |  |
| <10 years | 670 (18.8) | 1,282 (20.8) | 0.07 | 3,149 (23.3) | 3,149 (23.3) | 0.00 | 5,576 (25.6) | 3,496 (26.3) | 0.03 |
| 10-19 years | 300 (8.4) | 549 (8.9) |  | 1,475 (10.9) | 1,475 (10.9) |  | 1,692 (7.8) | 993 (7.5) |  |
| 20-29 years | 771 (21.6) | 1,286 (20.8) |  | 2,633 (19.5) | 2,633 (19.5) |  | 4,311 (19.8) | 2,608 (19.6) |  |
| 30-39 years | 1,037 (29.0) | 1,788 (29.0) |  | 3,427 (25.3) | 3,427 (25.3) |  | 5,692 (26.1) | 3,368 (25.4) |  |
| 40-49 years | 475 (13.3) | 797 (12.9) |  | 1,512 (11.2) | 1,512 (11.2) |  | 2,575 (11.8) | 1,568 (11.8) |  |
| 50-59 years | 231 (6.5) | 349 (5.7) |  | 880 (6.5) | 880 (6.5) |  | 1,346 (6.2) | 853 (6.4) |  |
| 60-69 years | 68 (1.9) | 89 (1.4) |  | 315 (2.3) | 315 (2.3) |  | 400 (1.8) | 261 (2.0) |  |
| 70+ years | 22 (0.6) | 36 (0.6) |  | 146 (1.1) | 146 (1.1) |  | 218 (1.0) | 141 (1.1) |  |
| **Sex** |  |  |  |  |  |  |  |  |  |
| Male | 1,769 (49.5) | 3,232 (52.3) | 0.06 | 7,717 (57.0) | 7,717 (57.0) | 0.00 | 12,678 (58.1) | 7,745 (58.3) | 0.00 |
| Female | 1,805 (50.5) | 2,944 (47.7) |  | 5,820 (43.0) | 5,820 (43.0) |  | 9,132 (41.9) | 5,543 (41.7) |  |
| **Nationality\*\*** |  |  |  |  |  |  |  |  |  |
| Bangladeshi | 74 (2.1) | 132 (2.1) | 0.07 | 443 (3.3) | 443 (3.3) | 0.00 | 762 (3.5) | 547 (4.1) | 0.06 |
| Egyptian | 224 (6.3) | 393 (6.4) |  | 897 (6.6) | 897 (6.6) |  | 1,249 (5.7) | 715 (5.4) |  |
| Filipino | 524 (14.7) | 890 (14.4) |  | 1,402 (10.4) | 1,402 (10.4) |  | 2,396 (11.0) | 1,389 (10.5) |  |
| Indian | 535 (15.0) | 1,007 (16.3) |  | 2,256 (16.7) | 2,256 (16.7) |  | 3,719 (17.1) | 2,306 (17.4) |  |
| Nepalese | 74 (2.1) | 132 (2.1) |  | 431 (3.2) | 431 (3.2) |  | 625 (2.9) | 363 (2.7) |  |
| Pakistani | 118 (3.3) | 221 (3.6) |  | 658 (4.9) | 658 (4.9) |  | 1,042 (4.8) | 633 (4.8) |  |
| Qatari | 866 (24.2) | 1,554 (25.2) |  | 3,364 (24.9) | 3,364 (24.9) |  | 5,117 (23.5) | 2,955 (22.2) |  |
| Sri Lankan | 42 (1.2) | 74 (1.2) |  | 262 (1.9) | 262 (1.9) |  | 444 (2.0) | 271 (2.0) |  |
| Sudanese | 212 (5.9) | 385 (6.2) |  | 789 (5.8) | 789 (5.8) |  | 1,273 (5.8) | 758 (5.7) |  |
| Other nationalities | 905 (25.3) | 1,388 (22.5) |  | 3,035 (22.4) | 3,035 (22.4) |  | 5,183 (23.8) | 3,351 (25.2) |  |

Abbreviations: IQR, interquartile range; PCR, polymerase chain reaction; SMD, standardized mean difference.
\*Cases and controls were matched one-to-two by sex, 10-year age group, nationality, and calendar week of PCR test.
†SMD is the difference in the mean of a covariate between groups divided by the pooled standard deviation. An SMD<0.1 indicates adequate matching.
‡Cases and controls were matched one-to-one by sex, 10-year age group, nationality, and calendar week of PCR test.
§Cases and controls were matched two-to-one by sex, 10-year age group, nationality, and calendar week of PCR test.
¶SMD is for the mean difference between groups divided by the pooled standard deviation.
\*\*Nationalities were chosen to represent the most populous groups in Qatar.

BNT162b2 effectiveness against symptomatic BA.1 infection was highest at 46.6% (95% confidence interval (CI): 33.4-57.2%) in the first three months after the second dose, but then declined to ∼10% or below thereafter (Figure 3A and Table 3). Effectiveness rapidly rebounded to 59.9% (95% CI: 51.2-67.0%) in the first month after the booster dose, but then declined to 40.5% (95% CI: 30.8-48.8%) in the second month and thereafter (only a small proportion of individuals were in the third or more months after the booster dose). A similar pattern was observed for mRNA-1273 effectiveness (Figure 3B and Table 4).

**Figure 3.**
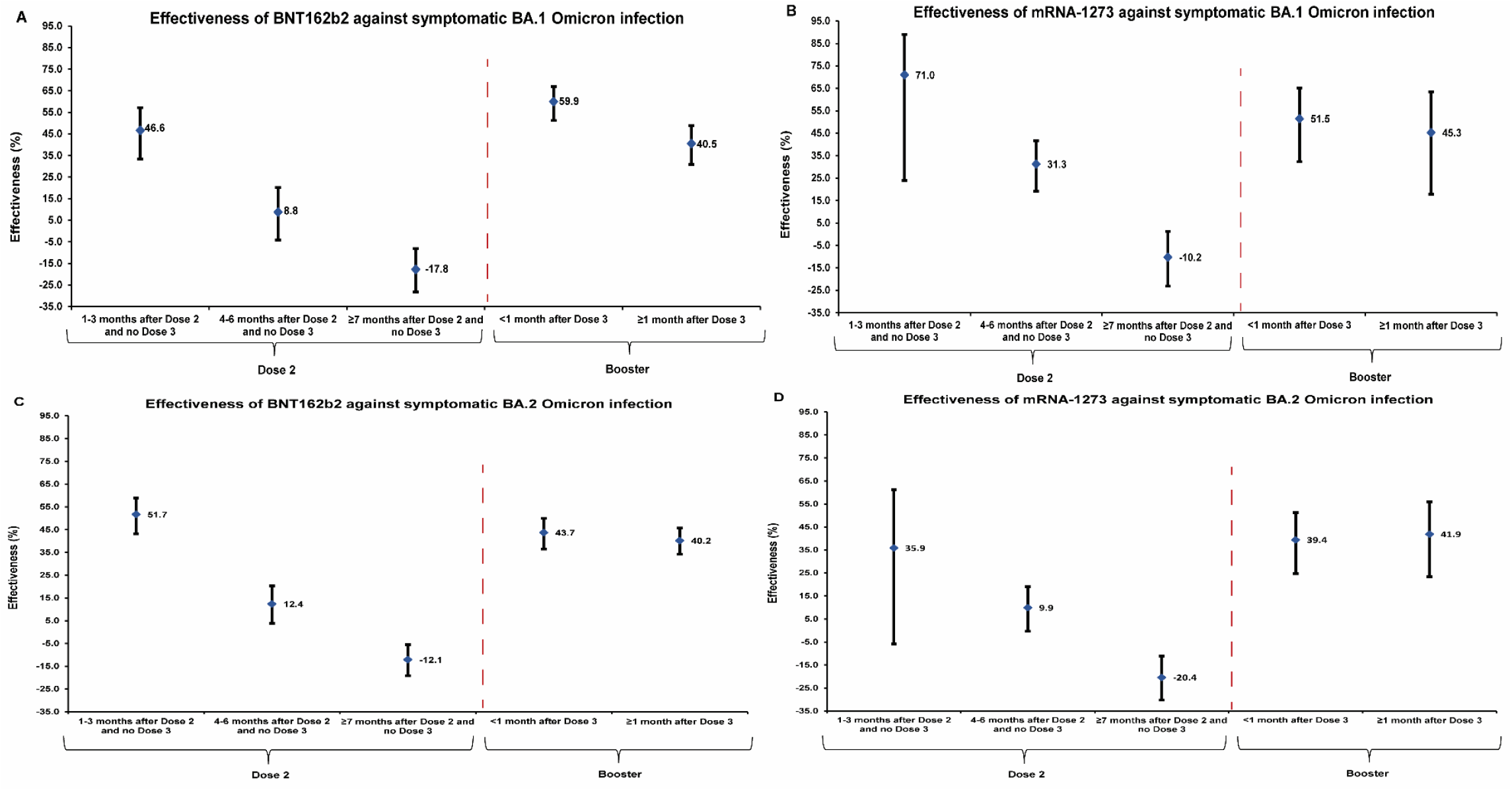
Effectiveness of the BNT162b2 and mRNA-1273 vaccines against symptomatic SARS-CoV-2 BA.1 Omicron infection (panels A and B, respectively) and symptomatic SARS-CoV-2 BA.2 Omicron infection (panels C and D, respectively). Data are presented as effectiveness point estimates. Error bars indicate the corresponding 95% confidence intervals.

**Table 3.** Effectiveness of the BNT162b2 vaccine against symptomatic SARS-CoV-2 BA.1 Omicron infection, BA.2 Omicron infection, and any Omicron infection*.

| Sub-studies<sup>†</sup> | Cases<br>(PCR-positive) |  | Controls<br>(PCR-negative) |  | Effectiveness in %<br>(95% CI)<sup>‡</sup> |
| --- | --- | --- | --- | --- | --- |
|  | Vaccinated | Unvaccinated | Vaccinated | Unvaccinated |  |
| Effectiveness against symptomatic BA.1 Omicron infection<sup>§</sup> |  |  |  |  |  |
| Dose 1 |  |  |  |  |  |
| 0-13 days after Dose 1 and no Dose 2 | 10 | 1,969 | 20 | 3,456 | 23.5 (-70.6 to 65.7) |
| ≥14 days after Dose 1 and no Dose 2 | 25 | 1,968 | 66 | 3,441 | 39.2 (2.3 to 62.1) |
| Dose 2 |  |  |  |  |  |
| 1-3 months after Dose 2 and no Dose 3 | 130 | 1,992 | 376 | 3,409 | 46.6 (33.4 to 57.2) |
| 4-6 months after Dose 2 and no Dose 3 | 502 | 2,004 | 941 | 3,506 | 8.8 (-4.1 to 20.1) |
| ≥7 months after Dose 2 and no Dose 3 | 3,570 | 2,060 | 6,007 | 3,947 | -17.8 (-28.2 to -8.2) |
| Dose 3 (booster dose) |  |  |  |  |  |
| <1 month after Dose 3 | 180 | 2,008 | 622 | 3,339 | 59.9 (51.2 to 67.0) |
| ≥1 month after Dose 3 | 483 | 2,031 | 1,145 | 3,441 | 40.5 (30.8 to 48.8) |
| Effectiveness against symptomatic BA.2 Omicron infection<sup>¶</sup> |  |  |  |  |  |
| Dose 1 |  |  |  |  |  |
| 0-13 days after Dose 1 and no Dose 2 | 32 | 5,744 | 34 | 5,742 | 5.9 (-52.5 to 41.9) |
| ≥14 days after Dose 1 and no Dose 2 | 64 | 5,774 | 99 | 5,739 | 36.1 (12.1 to 53.5) |
| Dose 2 |  |  |  |  |  |
| 1-3 months after Dose 2 and no Dose 3 | 263 | 5,964 | 496 | 5,731 | 51.7 (43.2 to 58.9) |
| 4-6 months after Dose 2 and no Dose 3 | 1,203 | 5,924 | 1,318 | 5,809 | 12.4 (3.8 to 20.3) |
| ≥7 months after Dose 2 and no Dose 3 | 8,003 | 5,840 | 7,762 | 6,081 | -12.1 (-19.1 to -5.5) |
| Dose 3 (booster dose) |  |  |  |  |  |
| <1 month after Dose 3 | 709 | 6,038 | 1,034 | 5,713 | 43.7 (36.5 to 50.0) |
| ≥1 month after Dose 3 | 1,580 | 6,211 | 2,029 | 5,762 | 40.2 (34.2 to 45.7) |
| Effectiveness against any symptomatic Omicron infection<sup>**</sup> |  |  |  |  |  |
| Dose 1 |  |  |  |  |  |
| 0-13 days after Dose 1 | 56 | 12,174 | 34 | 7,278 | 9.5 (-39.8 to 41.3) |
| ≥14 days after Dose 1 and no Dose 2 | 151 | 12,205 | 130 | 7,276 | 31.4 (12.5 to 46.3) |
| Dose 2 |  |  |  |  |  |
| 1-3 months after Dose 2 and no Dose 3 | 585 | 12,623 | 599 | 7,309 | 47.8 (40.8 to 53.9) |
| 4-6 months after Dose 2 and no Dose 3 | 2,479 | 12,590 | 1,605 | 7,333 | 16.3 (9.7 to 22.5) |
| ≥7 months after Dose 2 and no Dose 3 | 16,435 | 12,564 | 9,073 | 7,637 | -9.0 (-14.5 to -3.7) |
| Dose 3 (booster dose) |  |  |  |  |  |
| <1 month after Dose 3 | 1,326 | 12,777 | 1,300 | 7,275 | 49.5 (44.3 to 54.1) |
| ≥1 month after Dose 3 | 3,120 | 13,040 | 2,484 | 7,360 | 39.4 (34.4 to 44.0) |

Abbreviations: CI, confidence interval; PCR, polymerase chain reaction.
<sup>†</sup>A symptomatic infection was defined as a PCR-positive nasopharyngeal swab conducted because of clinical suspicion due to presence of symptoms compatible with a respiratory tract infection.
<sup>‡</sup>In each analysis for a specific time-since-vaccination stratum, we included only those vaccinated in this specific time-since-vaccination stratum and those unvaccinated. Only matched pairs of PCR-positive and PCR-negative persons, in which both members of the pair were either unvaccinated or fell within each time-since-vaccination stratum have been included in the corresponding vaccine effectiveness estimate. Thus, the number of cases (and controls) varied across time-since-vaccination analyses.
<sup>§</sup>Vaccine effectiveness was estimated using the test-negative, case-control study design<sup>11,12</sup>.
<sup>¶</sup>Cases and controls were matched one-to-two by sex, 10-year age group, nationality, and calendar week of PCR test.
<sup>\*\*</sup>Cases and controls were matched one-to-one by sex, 10-year age group, nationality, and calendar week of PCR test.
<sup>\*\*\*</sup>Cases and controls were matched two-to-one by sex, 10-year age group, nationality, and calendar week of PCR test.

**Table 4.**
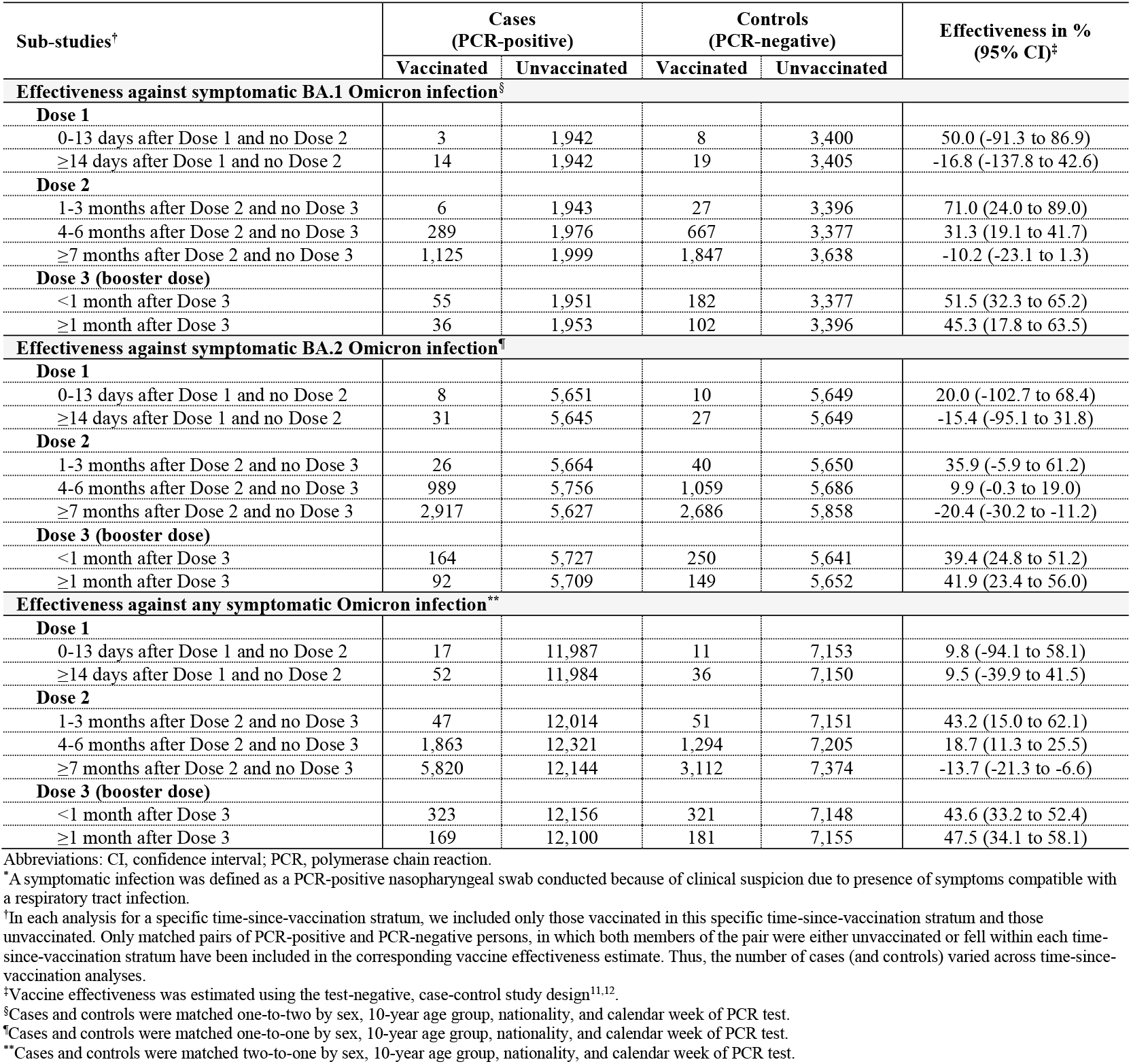
Effectiveness of the mRNA-1273 vaccine against symptomatic SARS-CoV-2 BA.1 Omicron infection, BA.2 Omicron infection, and any Omicron infection*.

| Sub-studies<sup>†</sup> | Cases<br>(PCR-positive) |  | Controls<br>(PCR-negative) |  | Effectiveness in %<br>(95% CI)<sup>‡</sup> |
| --- | --- | --- | --- | --- | --- |
|  | Vaccinated | Unvaccinated | Vaccinated | Unvaccinated |  |
| Effectiveness against symptomatic BA.1 Omicron infection<sup>§</sup> |  |  |  |  |  |
| Dose 1 |  |  |  |  |  |
| 0-13 days after Dose 1 and no Dose 2 | 3 | 1,942 | 8 | 3,400 | 50.0 (-91.3 to 86.9) |
| ≥14 days after Dose 1 and no Dose 2 | 14 | 1,942 | 19 | 3,405 | -16.8 (-137.8 to 42.6) |
| Dose 2 |  |  |  |  |  |
| 1-3 months after Dose 2 and no Dose 3 | 6 | 1,943 | 27 | 3,396 | 71.0 (24.0 to 89.0) |
| 4-6 months after Dose 2 and no Dose 3 | 289 | 1,976 | 667 | 3,377 | 31.3 (19.1 to 41.7) |
| ≥7 months after Dose 2 and no Dose 3 | 1,125 | 1,999 | 1,847 | 3,638 | -10.2 (-23.1 to 1.3) |
| Dose 3 (booster dose) |  |  |  |  |  |
| <1 month after Dose 3 | 55 | 1,951 | 182 | 3,377 | 51.5 (32.3 to 65.2) |
| ≥1 month after Dose 3 | 36 | 1,953 | 102 | 3,396 | 45.3 (17.8 to 63.5) |
| Effectiveness against symptomatic BA.2 Omicron infection<sup>¶</sup> |  |  |  |  |  |
| Dose 1 |  |  |  |  |  |
| 0-13 days after Dose 1 and no Dose 2 | 8 | 5,651 | 10 | 5,649 | 20.0 (-102.7 to 68.4) |
| ≥14 days after Dose 1 and no Dose 2 | 31 | 5,645 | 27 | 5,649 | -15.4 (-95.1 to 31.8) |
| Dose 2 |  |  |  |  |  |
| 1-3 months after Dose 2 and no Dose 3 | 26 | 5,664 | 40 | 5,650 | 35.9 (-5.9 to 61.2) |
| 4-6 months after Dose 2 and no Dose 3 | 989 | 5,756 | 1,059 | 5,686 | 9.9 (-0.3 to 19.0) |
| ≥7 months after Dose 2 and no Dose 3 | 2,917 | 5,627 | 2,686 | 5,858 | -20.4 (-30.2 to -11.2) |
| Dose 3 (booster dose) |  |  |  |  |  |
| <1 month after Dose 3 | 164 | 5,727 | 250 | 5,641 | 39.4 (24.8 to 51.2) |
| ≥1 month after Dose 3 | 92 | 5,709 | 149 | 5,652 | 41.9 (23.4 to 56.0) |
| Effectiveness against any symptomatic Omicron infection<sup>**</sup> |  |  |  |  |  |
| Dose 1 |  |  |  |  |  |
| 0-13 days after Dose 1 and no Dose 2 | 17 | 11,987 | 11 | 7,153 | 9.8 (-94.1 to 58.1) |
| ≥14 days after Dose 1 and no Dose 2 | 52 | 11,984 | 36 | 7,150 | 9.5 (-39.9 to 41.5) |
| Dose 2 |  |  |  |  |  |
| 1-3 months after Dose 2 and no Dose 3 | 47 | 12,014 | 51 | 7,151 | 43.2 (15.0 to 62.1) |
| 4-6 months after Dose 2 and no Dose 3 | 1,863 | 12,321 | 1,294 | 7,205 | 18.7 (11.3 to 25.5) |
| ≥7 months after Dose 2 and no Dose 3 | 5,820 | 12,144 | 3,112 | 7,374 | -13.7 (-21.3 to -6.6) |
| Dose 3 (booster dose) |  |  |  |  |  |
| <1 month after Dose 3 | 323 | 12,156 | 321 | 7,148 | 43.6 (33.2 to 52.4) |
| ≥1 month after Dose 3 | 169 | 12,100 | 181 | 7,155 | 47.5 (34.1 to 58.1) |

Abbreviations: CI, confidence interval; PCR, polymerase chain reaction.
<sup>†</sup>A symptomatic infection was defined as a PCR-positive nasopharyngeal swab conducted because of clinical suspicion due to presence of symptoms compatible with a respiratory tract infection.
<sup>‡</sup>In each analysis for a specific time-since-vaccination stratum, we included only those vaccinated in this specific time-since-vaccination stratum and those unvaccinated. Only matched pairs of PCR-positive and PCR-negative persons, in which both members of the pair were either unvaccinated or fell within each time-since-vaccination stratum have been included in the corresponding vaccine effectiveness estimate. Thus, the number of cases (and controls) varied across time-since-vaccination analyses.
<sup>§</sup>Vaccine effectiveness was estimated using the test-negative, case-control study design<sup>11,12</sup>.
<sup>¶</sup>Cases and controls were matched one-to-two by sex, 10-year age group, nationality, and calendar week of PCR test.
<sup>\*\*</sup>Cases and controls were matched one-to-one by sex, 10-year age group, nationality, and calendar week of PCR test.
<sup>\*\*\*</sup>Cases and controls were matched two-to-one by sex, 10-year age group, nationality, and calendar week of PCR test.

BNT162b2 effectiveness against symptomatic BA.2 infection was highest at 51.7% (95% CI: 43.2-58.9%) in the first three months after the second dose, but then declined to ∼10% or below thereafter (Figure 3C and Table 3). Effectiveness rapidly rebounded to 43.7% (95% CI: 36.5-50.0%) in the first month after the booster dose, but then declined to 40.2% (95% CI: 34.2-45.7%) in the second month and thereafter. A similar pattern was observed for mRNA-1273 effectiveness (Figure 3D and Table 4).

BNT162b2 effectiveness against any symptomatic Omicron infection, regardless of subvariant, was highest at 47.8% (95% CI: 40.8-53.9%) in the first three months after the second dose, but then declined to ∼15% or below thereafter (Figure 4A and Table 3). Effectiveness rapidly rebounded to 49.5% (95% CI: 44.3-54.1%) in the first month after the booster dose, but then declined to 39.4% (95% CI: 34.4-44.0%) in the second month and thereafter. A similar pattern was observed for mRNA-1273 effectiveness (Figure 4B and Table 4).

**Figure 4.**
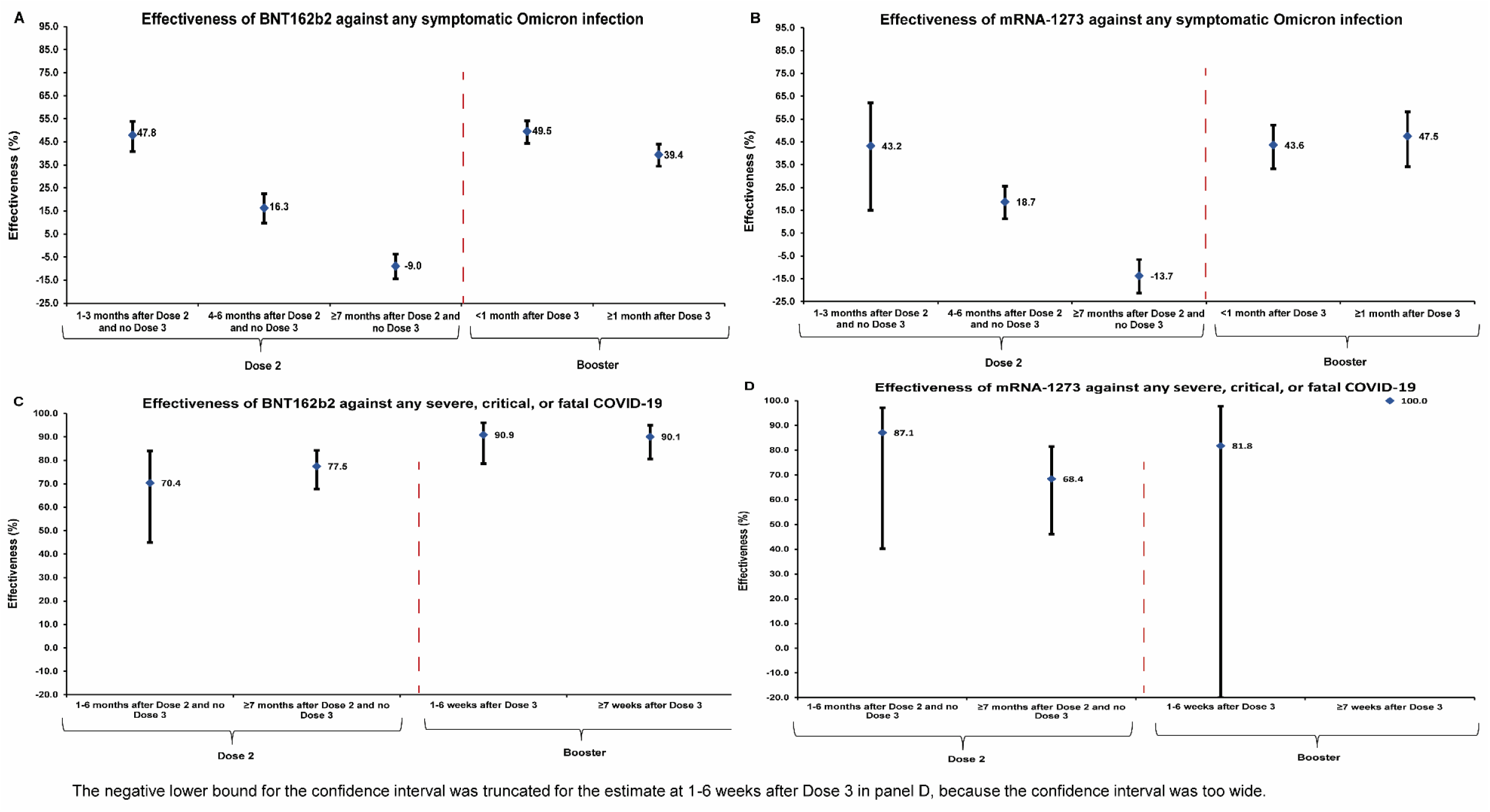
Effectiveness of the BNT162b2 and mRNA-1273 vaccines against any symptomatic SARS-CoV-2 Omicron infection regardless of subvariant (panels A and B, respectively) and against any severe^9^, critical^9^, or fatal^10^ COVID-19 due to an Omicron infection (panels C and D, respectively). Data are presented as effectiveness point estimates. Error bars indicate the corresponding 95% confidence intervals.

Effectiveness against any severe, critical, or fatal COVID-19 due to an Omicron infection, regardless of subvariant, was in the range of 70-80% at any time after the second dose for both the BNT162b2 and mRNA-1273 vaccines (Figures 4C and 4D and Table 5). However, BNT162b2 effectiveness against any severe, critical, or fatal COVID-19 after the booster dose was greater than 90%. 95% CIs around estimates of mRNA-1273 effectiveness against any severe, critical, or fatal COVID-19 after the booster dose lacked adequate statistical precision— there were too few hospitalized COVID-19 cases among mRNA-1273 vaccinated persons (Table 5).

**Table 5.** Effectiveness of the BNT162b2 and mRNA-1273 vaccines against any severe^9^, critical^9^, or fatal^10^ COVID-19.

| Sub-studies* | BNT162b2 |  |  |  |  | mRNA-1273 |  |  |  |  |
| --- | --- | --- | --- | --- | --- | --- | --- | --- | --- | --- |
|  | Cases†<br>(Severe, critical, or<br>fatal disease)‡ |  | Controls†<br>(PCR-negative) |  | Effectiveness in %<br>(95% CI)§ | Cases†<br>(Severe, critical, or<br>fatal disease)‡ |  | Controls†<br>(PCR-negative) |  | Effectiveness in %<br>(95% CI)§ |
|  | Vaccinated |  | Vaccinated |  |  | Vaccinated |  | Vaccinated |  |  |
|  | Yes | No | Yes | No |  | Yes | No | Yes | No |  |
| Dose 1 |  |  |  |  |  |  |  |  |  |  |
| Dose 1 and no Dose 2 | 2 | 111 | 8 | 290 | 40.9 (-199.1 to 88.3) | 0 | 110 | 3 | 287 | 100.0 (Omitted)¶ |
| Dose 2 |  |  |  |  |  |  |  |  |  |  |
| 1-6 months after Dose 2 and no Dose 3 | 14 | 123 | 108 | 261 | 70.4 (45.0 to 84.0) | 2 | 113 | 34 | 272 | 87.1 (40.2 to 97.2) |
| ≥7 months after Dose 2 and no Dose 3 | 76 | 143 | 461 | 218 | 77.5 (67.8 to 84.3) | 23 | 126 | 148 | 264 | 68.4 (46.1 to 81.5) |
| Dose 3 (booster dose) |  |  |  |  |  |  |  |  |  |  |
| 1-6 weeks after Dose 3 | 8 | 125 | 143 | 257 | 90.9 (78.6 to 96.1) | 1 | 110 | 18 | 280 | 81.8 (-49.5 to 97.8) |
| ≥7 weeks after Dose 3 | 12 | 134 | 197 | 254 | 90.1 (80.6 to 95.0) | 0 | 110 | 3 | 287 | 100.0 (Omitted)¶ |

Abbreviations: CI, confidence interval; PCR, polymerase chain reaction.
<sup>\*</sup>In each analysis for a specific time-since-vaccination stratum, we included only those vaccinated in this specific time-since-vaccination stratum and those unvaccinated. Only matched pairs of PCR-positive and PCR-negative persons, in which both members of the pair were either unvaccinated or fell within each time-since-vaccination stratum have been included in the corresponding vaccine effectiveness estimate. Thus, the number of cases (and controls) varied across time-since-vaccination analyses.
<sup>†</sup>Cases and controls were matched one-to-five by sex, 10-year age group, nationality, and calendar week of PCR test.
<sup>‡</sup>Severity<sup>9</sup>, criticality<sup>9</sup>, and fatality<sup>10</sup> were defined as per World Health Organization guidelines.
<sup>§</sup>Vaccine effectiveness was estimated using the test-negative, case-control study design<sup>11,12</sup>.
<sup>¶</sup>Confidence interval could not be estimated using conditional logistic regression because of zero events among those vaccinated.

### Additional analyses

Sensitivity analyses adjusting for documented prior infection and healthcare worker status yielded similar findings to the main analyses (Supplementary Tables 1 and 2). Sensitivity analyses to assess the impact of excluding children <12 years of age (Supplementary Tables 3 and 4), or individuals <20 years of age (Supplementary Tables 5 and 6), also yielded similar findings to the main analyses.

## Discussion

No discernable differences were observed in the duration of mRNA vaccine protection against BA.1 versus BA.2 symptomatic infection. For each of these subvariants, vaccine effectiveness against symptomatic infection was ∼50% in the first three months after the second dose, but declined to negligible levels thereafter. Effectiveness rapidly rebounded after the booster dose to reach similar levels to those seen right after the second dose, but appeared to wane again starting from the second month after the booster dose. There were also no discernable differences in effectiveness of BNT162b2 vaccine versus mRNA-1273 vaccine.

Despite only moderate and rapidly waning protection against symptomatic infection, mRNA vaccine effectiveness against COVID-19 hospitalization and death due to Omicron infections was strong at greater than 70% after the second dose. It was also higher after the booster dose at greater than 90%. These findings support the durability of vaccine protection against COVID-19 hospitalization and death for at least several months after receiving the second dose,^13,14^ but also demonstrate the importance of booster vaccination in achieving robust protection against any hospitalization and death due to Omicron infections. These findings suggest the need to consider rapid implementation of booster vaccination campaigns coincident with the emergence of a new wave or variant, at least to those most vulnerable to COVID-19 hospitalization and death.

This study has limitations. With the lower severity of Omicron infections^20^ and the young population of Qatar^15,21^, case numbers were insufficient to estimate the duration of protection against COVID-19 hospitalization and death for each subvariant separately. BA.1 and BA.2 ascertainment was based on proxy criteria, presence or absence of an S-gene “target failure” using the TaqPath PCR assay (Methods), but this method of ascertainment is well established not only for Omicron subvariants, but also for other variants such as Alpha^22-24^. Some Omicron infections may have been misclassified Delta infections, but this is not likely, as Delta incidence was limited during the study duration (Methods).

Vaccine protection was assessed for only several months after the second dose, and only several weeks after the booster dose. Longer-term protection against symptomatic infection and COVID-19 hospitalization and death remain uncertain. Vaccine effectiveness reached small but statistically significant negative values at 7 months or more after the second dose. Negative estimated effectiveness likely reflects an effect of bias and not true negative biological effectiveness. This bias may have risen from vaccinated persons having a higher social contact rate or adhering less to safety measures than unvaccinated persons^25-27^. With the high vaccine coverage among adults in Qatar (>85%)^13^, this bias may have also risen because the reference group of unvaccinated individuals included mainly children or young persons; therefore, it may not be representative of the wider population. However, sensitivity analyses excluding children and young persons confirmed the same study findings (Supplementary Tables 3-6).

Unvaccinated adults are a small minority that may not be truly immune-naïve due to undocumented prior SARS-CoV-2 infections, especially now that we are two years into this pandemic. Bias due to depletion of the susceptible population may lead to underestimation of vaccine effectiveness^28^, even in the test-negative, case-control, study design, which is less prone to effect of this bias^13^.

While matching was done for sex, age, and nationality, this was not possible for other factors, such as comorbidities, as such data are not available. However, matching by these factors provided demonstrable control of bias in studies of different epidemiologic designs and that used control groups in Qatar^13,14,29-31^. Effectiveness was assessed using an observational, test-negative, case-control, study design^11,12^, rather than a design in which cohorts of vaccinated and unvaccinated individuals were followed up. However, the cohort study design applied earlier to the same population of Qatar yielded findings similar to those of the test-negative case-control design^30,32,33^, supporting the validity of this standard approach in assessing vaccine effectiveness^11,12,30,34^. Moreover, our recent study of the effectiveness of booster vaccination against any symptomatic Omicron infection, relative to that of the primary series, used a cohort study design^5^ and its results are consistent with results generated in the present study using the test-negative, case-control study design.

Nonetheless, one cannot exclude the possibility that in real-world data, bias could arise in unexpected ways, or from unknown sources, such as subtle differences in test-seeking behavior or changes in the pattern of testing with introduction of other testing modalities, such as rapid antigen testing (RAT). For example, with the large Omicron wave in Qatar, use of RAT was expanded to supplement PCR testing starting from January 5, 2022. However, RAT was broadly implemented in the population and probably did not differentially affect PCR testing to introduce bias. With the small proportion of Qatar’s population ≥50 years of age^15,35^, our findings may not be generalizable to other countries in which elderly citizens constitute a larger proportion of the total population.

Notwithstanding these limitations, consistent findings were reached, indicating rapid waning of vaccine protection against symptomatic Omicron infection that are consistent with findings of other studies for effectiveness against Omicron infection (with no BA.1/BA.2 subvariant specified)^36-41^. Moreover, with the mass scale of PCR testing in Qatar^13^, the likelihood of bias is perhaps minimized. Extensive sensitivity and additional analyses were conducted to investigate effects of potential bias in our earlier studies for the BNT162b2^13^ and mRNA-1273^14^ vaccines, which used the same methodology used here. These included different adjustments in the analysis, different approaches for factoring prior infection in the analysis, and different study inclusion and exclusion criteria to investigate whether effectiveness estimates could have been biased^13,14^. These analyses showed consistent findings^13,14^.

In conclusion, mRNA vaccines provide only moderate protection against symptomatic BA.1 and BA.2 Omicron infections, with no discernable differences in protection against either the BA.1 or BA.2 subvariants. Protection also wanes rapidly to negligible levels, starting four months after the second dose. Vaccine protection rebounds after booster vaccination, but also appears to wane thereafter. Meanwhile, vaccine protection against COVID-19 hospitalization and death is strong and durable after the second dose, and is most robust after a booster dose.

## Data Availability

The dataset of this study is a property of the Qatar Ministry of Public Health that was provided to the researchers through a restricted-access agreement that prevents sharing the dataset with a third party or publicly for preservation of confidentiality of patient data. Access to this dataset is at the discretion of the Qatar Ministry of Public Health. Access to the dataset can be considered through a direct application for data access to Her Excellency the Minister of Public Health (https://www.moph.gov.qa/english/Pages/default.aspx). Aggregate data are available within the manuscript and its supplementary information.

## Acknowledgements

We acknowledge the many dedicated individuals at Hamad Medical Corporation, the Ministry of Public Health, the Primary Health Care Corporation, the Qatar Biobank, Sidra Medicine, and Weill Cornell Medicine – Qatar for their diligent efforts and contributions to make this study possible. The authors are grateful for support from the Biomedical Research Program and the Biostatistics, Epidemiology, and Biomathematics Research Core, both at Weill Cornell Medicine-Qatar, as well as for support provided by the Ministry of Public Health, Hamad Medical Corporation, and Sidra Medicine. The authors are also grateful for the Qatar Genome Programme and Qatar University Biomedical Research Center for institutional support for the reagents needed for the viral genome sequencing. Statements made herein are solely the responsibility of the authors. The funders of the study had no role in study design, data collection, data analysis, data interpretation, or writing of the article.

## Author contributions

HC co-designed the study, performed the statistical analyses, and co-wrote the first draft of the article. LJA conceived and co-designed the study, led the statistical analyses, and co-wrote the first draft of the article. PT and MRH conducted the multiplex, RT-qPCR variant screening and viral genome sequencing. HY, HAK, and MKS conducted viral genome sequencing. All authors contributed to data collection and acquisition, database development, discussion and interpretation of the results, and to the writing of the manuscript. All authors have read and approved the final manuscript.

## Methods Oversight

Hamad Medical Corporation and Weill Cornell Medicine-Qatar Institutional Review Boards approved this retrospective study with waiver of informed consent. The study was reported following the Strengthening the Reporting of Observational studies in Epidemiology (STROBE) guidelines. The STROBE checklist is found in Supplementary Table 7.

### Study population and data sources

This study was conducted in the resident population of Qatar, applying methodology that was developed earlier to assess duration of protection of the BNT162b2^13^ and mRNA-1273^14^ coronavirus disease 2019 (COVID-19) vaccines in the same population during earlier acute respiratory syndrome coronavirus 2 (SARS-CoV-2) infection waves. COVID-19 laboratory testing, vaccination, clinical infection data, and demographic details were extracted from the national, federated SARS-CoV-2 databases that include, with no missing information, all polymerase chain reaction (PCR) testing, COVID-19 vaccinations, and COVID-19 hospitalizations and deaths in Qatar since the start of the pandemic.

Every PCR test conducted in Qatar is categorized based on symptoms and the reason for testing. Qatar has young, international demographics. Only 9% of Qatar’s population is ≥50 years of age and 89% are international expatriates from over 150 countries^15,35^. The vast majority of individuals were vaccinated in Qatar, but if vaccinated elsewhere, those vaccinations were still registered in the health system at the port of entry upon arrival in Qatar.

### Study design

Vaccine effectiveness against symptomatic SARS-CoV-2 Omicron (B.1.1.529)^1^ infection during the large Omicron wave in Qatar, between December 23, 2021 and February 28, 2022, was estimated using the test-negative, case-control study design, a standard design for assessing vaccine effectiveness^11,12,30,34^. A symptomatic Omicron infection was defined as a nasopharyngeal PCR-positive swab collected during the Omicron wave because of clinical suspicion of infection, i.e., symptoms indicative of a respiratory tract infection. Cases (BA.1, BA.2, or any-Omicron-subvariant infected persons) and controls (uninfected persons) were exact-matched by sex, 10-year age group, nationality, and calendar week of PCR test. The ratio of matching in each analysis was determined based on available cases and controls (Figure 2). Matching was implemented to control for established differences in the risk of exposure to SARS-CoV-2 infection in Qatar^15-19^.

Only the first PCR-positive test during the study was included for each case, whereas all PCR-negative tests during the study were included for each control. Controls included individuals with no record of a positive PCR or rapid-antigen test (RAT) during the study period. Only PCR tests conducted because of clinical suspicion of infection, i.e., symptoms indicative of a respiratory tract infection, were included in the analysis for cases and controls. All persons who received a vaccine other than BNT162b2 or mRNA-1273, or who received a different mix of vaccines, were excluded. These inclusion and exclusion criteria were implemented to minimize different types of potential bias based on earlier analyses in the same population^13,14^. Every case (or control) that met the inclusion criteria and that could be matched to a control (case) was included in the analysis. COVID-19 vaccination status was ascertained at the time of the PCR test.

Vaccine effectiveness was also estimated against any severe, critical, or fatal COVID-19 infection due to Omicron, using the same methodology. Classification of COVID-19 case severity (acute-care hospitalizations)^9^, criticality (intensive-care-unit hospitalizations)^9^, and fatality^10^ followed World Health Organization (WHO) guidelines, and assessments were made by trained medical personnel using individual chart reviews (detailed description below). Each person who had a PCR-positive test result and COVID-19 hospital admission was subject to an infection severity assessment every three days until discharge or death, regardless of the hospital stay length or the time between the PCR-positive test and the final disease outcome. Individuals who progressed to severe^9^, critical^9^, or fatal^10^ COVID-19 between the PCR-positive test result and the end of the study were classified based on their worst outcome, starting with death, followed by critical disease, and then severe disease.

### COVID-19 severity, criticality, and fatality classification

WHO defines severe COVID-19 as a SARS-CoV-2 infected individual with “oxygen saturation of <90% on room air, and/or respiratory rate of >30 breaths/minute in adults and children >5 years old (or ≥60 breaths/minute in children <2 months old or ≥50 breaths/minute in children 2– 11 months old or ≥40 breaths/minute in children 1–5 years old), and/or signs of severe respiratory distress (accessory muscle use and inability to complete full sentences, and, in children, very severe chest wall indrawing, grunting, central cyanosis, or presence of any other general danger signs)”^9^. Detailed criteria are in the WHO technical report^9^.

Critical COVID-19 is defined as a SARS-CoV-2 infected individual with “acute respiratory distress syndrome, sepsis, septic shock, or other conditions that would normally require the provision of life sustaining therapies such as mechanical ventilation (invasive or non-invasive) or vasopressor therapy”^9^. Detailed criteria are in the WHO technical report^9^.

COVID-19 death is defined as “a death resulting from a clinically compatible illness, in a probable or confirmed COVID-19 case, unless there is a clear alternative cause of death that cannot be related to COVID-19 disease (e.g., trauma). There should be no period of complete recovery from COVID-19 between illness and death. A death due to COVID-19 may not be attributed to another disease (e.g., cancer) and should be counted independently of preexisting conditions that are suspected of triggering a severe course of COVID-19”. Detailed criteria are in the WHO technical report^10^.

### Laboratory methods and subvariant ascertainment

#### Real-time reverse-transcription polymerase chain reaction testing

Nasopharyngeal and/or oropharyngeal swabs were collected for PCR testing and placed in Universal Transport Medium (UTM). Aliquots of UTM were: 1) extracted on KingFisher Flex (Thermo Fisher Scientific, USA), MGISP-960 (MGI, China), or ExiPrep 96 Lite (Bioneer, South Korea) followed by testing with real-time reverse-transcription PCR (RT-qPCR) using TaqPath COVID-19 Combo Kits (Thermo Fisher Scientific, USA) on an ABI 7500 FAST (Thermo Fisher Scientific, USA); 2) tested directly on the Cepheid GeneXpert system using the Xpert Xpress SARS-CoV-2 (Cepheid, USA); or 3) loaded directly into a Roche cobas 6800 system and assayed with the cobas SARS-CoV-2 Test (Roche, Switzerland). The first assay targets the viral S, N, and ORF1ab gene regions. The second targets the viral N and E-gene regions, and the third targets the ORF1ab and E-gene regions.

All PCR testing was conducted at the Hamad Medical Corporation Central Laboratory or at the Sidra Medicine Laboratory, following standardized protocols.

#### Classification of infections by subvariant

Surveillance for SARS-CoV-2 variants in Qatar is mainly based on viral genome sequencing and multiplex RT-qPCR variant screening^42^ of random positive clinical samples^2,13,30,32,43,44^, complemented by deep sequencing of wastewater samples^2,45^.

A total of 315 random SARS-CoV-2-positive specimens collected between December 19, 2021 and January 22, 2022 were viral whole-genome sequenced on a Nanopore GridION sequencing device. Of these, 300 (95.2%) were confirmed as Omicron infections and 15 (4.8%) as Delta (B.1.617.2)^1^ infections^2,4,5^. Of 286 Omicron infections with confirmed subvariant status, 68 (23.8%) were BA.1 cases and 218 (76.2%) were BA.2 cases. No Delta case was detected in sequencing after January 8, 2022, nor were other variants.

Additionally, a total of 1,315 random SARS-CoV-2-positive specimens collected between December 22, 2021 and January 1, 2022 were RT-qPCR genotyped. The RT-qPCR genotyping identified 1 B.1.617.2-like Delta case, 366 BA.1-like Omicron cases, 898 BA.2-like Omicron cases, and 50 were undetermined cases where the genotype could not be assigned.

The accuracy of the RT-qPCR genotyping was verified against either Sanger sequencing of the receptor-binding domain (RBD) of SARS-CoV-2 surface glycoprotein (S) gene, or by viral whole-genome sequencing on a Nanopore GridION sequencing device. From 147 random SARS-CoV-2-positive specimens all collected in December of 2021, RT-qPCR genotyping was able to assign a genotype in 129 samples. The agreement between RT-qPCR genotyping and sequencing was 100% for Delta (n=82), 100% for Omicron BA.1 (n=18), and 93% for Omicron BA.2 (27 of 29 were correctly assigned to BA.2 and remaining 2 specimens genotyped as BA.2 were B.1.617.2 by sequencing). Of the remaining 18 specimens: 10 failed PCR amplification and sequencing, 8 could not be assigned a genotype by RT-qPCR (4 of 8 were B.1.617.2 by sequencing, and the remaining 4 failed sequencing). All the variant RT-qPCR genotyping was conducted at the Sidra Medicine Laboratory following standardized protocols.

The large Omicron-wave exponential-growth phase in Qatar started on December 19, 2021 and peaked in mid-January, 2022^2-5^. The study duration coincided with the intense Omicron wave where Delta incidence was limited. Accordingly, any PCR-positive test during the study duration, between December 23, 2021 and February 28, 2022, was assumed to be an Omicron infection. Of note that the study duration started on December 23, 2021, and not on December 19, 2021, to minimize the occurrence of residual Delta incidence during the first few days of the Omicron wave.

Informed by the viral genome sequencing and the RT-qPCR genotyping, a SARS-CoV-2 infection with the BA.1 subvariant was proxied as an S-gene “target failure” (SGTF) case using the TaqPath COVID-19 Combo Kit (Thermo Fisher Scientific, USA^46^) that tests for the S-gene and is affected by the del69/70 mutation in the S-gene^22^. A SARS-CoV-2 infection with the BA.2 subvariant was proxied as a non-SGTF case using this TaqPath Kit.

### Statistical analysis

Study samples were described using frequency distributions and measures of central tendency. Groups were compared using standardized mean differences (SMDs), defined as the difference in the mean of a covariate between groups, divided by the pooled standard deviation. SMD <0.1 indicated adequate matching^47^. The odds ratio (and 95% confidence interval (CI)), comparing odds of vaccination among cases to that among controls, was estimated using conditional logistic regression factoring the matching in the study design. This analytical approach was implemented to reduce potential bias due to variation in epidemic phase^11,48^, gradual vaccination roll-out^11,48^, and other confounders^15,17-19,49,50^. CIs did not factor multiplicity and should not be used to infer definitive differences between study groups. Interactions were not examined. Vaccine effectiveness at different time frames and its associated 95% CI were then estimated using^11,12^:

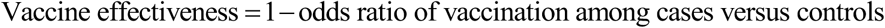

Since we used a test-negative study design, some persons were tested, PCR-positive or PCR-negative, after one vaccine dose, but before the next vaccine dose. This allowed us to estimate effectiveness after each dose. In each time-since-vaccination stratum, for first, second, and third doses, we analyzed only those vaccinated in this specific time-since-vaccination stratum and those unvaccinated (our reference group). Accordingly, the sample size for cases (and controls) varied in the different time-since-vaccination analyses. Effectiveness was estimated by one or more months in which one month was defined as 30 days, or by one or more weeks where one week was defined as 7 days.

Sensitivity analyses were conducted to assess the impact on effectiveness estimates of adjusting for documented prior infection and healthcare worker status in conditional logistic regression. With the majority of those unvaccinated being children or young persons, and therefore not necessarily representative of total population demographics, additional analyses were conducted to assess the impact of excluding children <12 years of age and individuals <20 years of age on effectiveness estimates. Statistical analyses were conducted in STATA/SE version 17.0^51^.

## Supplementary Appendix

**Supplementary Table 1.**
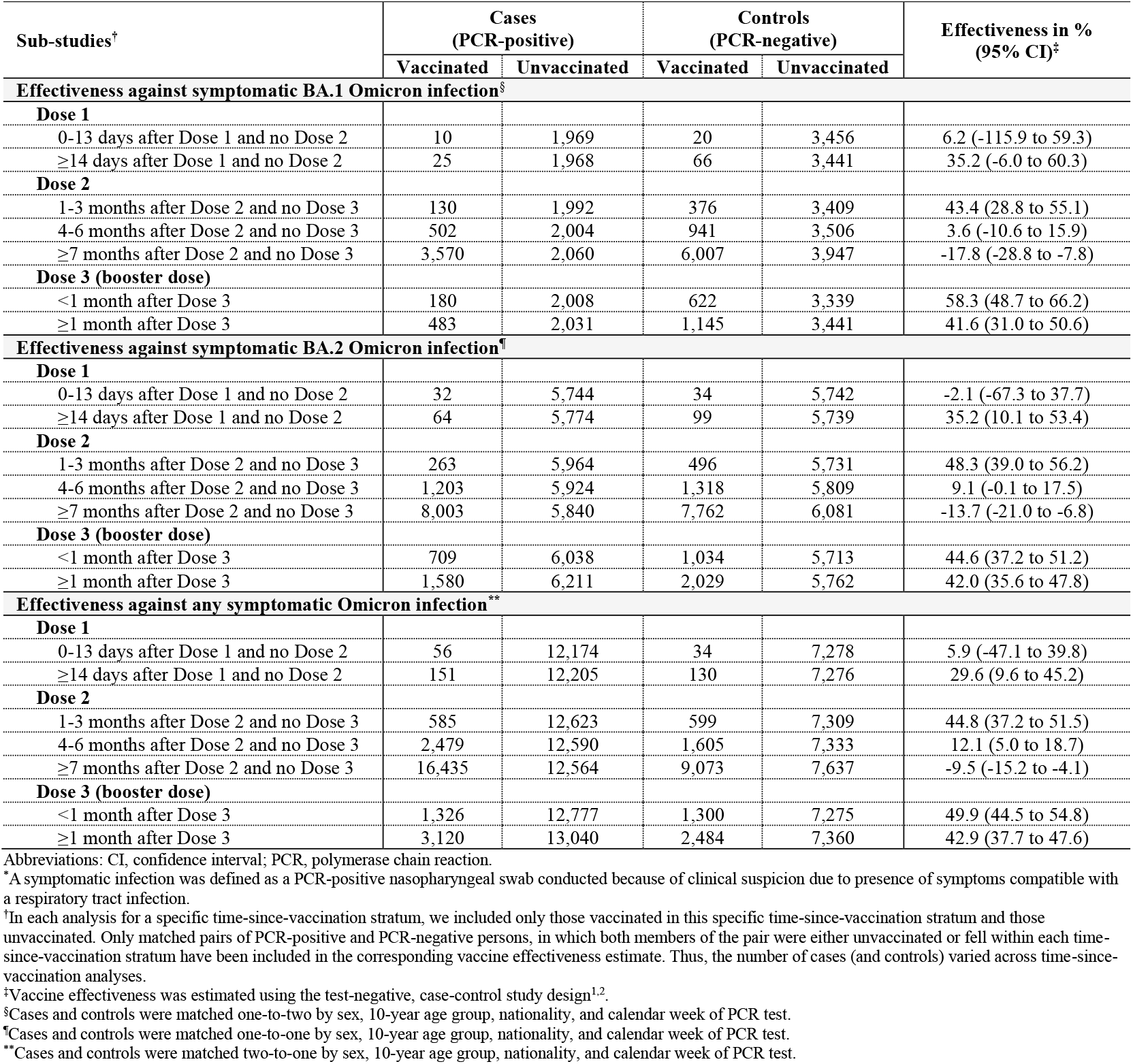
Sensitivity analysis for the effectiveness of the BNT162b2 vaccine against symptomatic SARS-CoV-2 BA.1 Omicron infection, BA.2 Omicron infection, and any Omicron infection*, adjusting for documented prior infection and health worker status in the conditional logistic regression.

| Sub-studies<sup>†</sup> | Cases<br>(PCR-positive) |  | Controls<br>(PCR-negative) |  | Effectiveness in %<br>(95% CI)<sup>‡</sup> |
| --- | --- | --- | --- | --- | --- |
|  | Vaccinated | Unvaccinated | Vaccinated | Unvaccinated |  |
| Effectiveness against symptomatic BA.1 Omicron infection<sup>§</sup> |  |  |  |  |  |
| Dose 1 |  |  |  |  |  |
| 0-13 days after Dose 1 and no Dose 2 | 10 | 1,969 | 20 | 3,456 | 6.2 (-115.9 to 59.3) |
| ≥14 days after Dose 1 and no Dose 2 | 25 | 1,968 | 66 | 3,441 | 35.2 (-6.0 to 60.3) |
| Dose 2 |  |  |  |  |  |
| 1-3 months after Dose 2 and no Dose 3 | 130 | 1,992 | 376 | 3,409 | 43.4 (28.8 to 55.1) |
| 4-6 months after Dose 2 and no Dose 3 | 502 | 2,004 | 941 | 3,506 | 3.6 (-10.6 to 15.9) |
| ≥7 months after Dose 2 and no Dose 3 | 3,570 | 2,060 | 6,007 | 3,947 | -17.8 (-28.8 to -7.8) |
| Dose 3 (booster dose) |  |  |  |  |  |
| <1 month after Dose 3 | 180 | 2,008 | 622 | 3,339 | 58.3 (48.7 to 66.2) |
| ≥1 month after Dose 3 | 483 | 2,031 | 1,145 | 3,441 | 41.6 (31.0 to 50.6) |
| Effectiveness against symptomatic BA.2 Omicron infection<sup>§</sup> |  |  |  |  |  |
| Dose 1 |  |  |  |  |  |
| 0-13 days after Dose 1 and no Dose 2 | 32 | 5,744 | 34 | 5,742 | -2.1 (-67.3 to 37.7) |
| ≥14 days after Dose 1 and no Dose 2 | 64 | 5,774 | 99 | 5,739 | 35.2 (10.1 to 53.4) |
| Dose 2 |  |  |  |  |  |
| 1-3 months after Dose 2 and no Dose 3 | 263 | 5,964 | 496 | 5,731 | 48.3 (39.0 to 56.2) |
| 4-6 months after Dose 2 and no Dose 3 | 1,203 | 5,924 | 1,318 | 5,809 | 9.1 (-0.1 to 17.5) |
| ≥7 months after Dose 2 and no Dose 3 | 8,003 | 5,840 | 7,762 | 6,081 | -13.7 (-21.0 to -6.8) |
| Dose 3 (booster dose) |  |  |  |  |  |
| <1 month after Dose 3 | 709 | 6,038 | 1,034 | 5,713 | 44.6 (37.2 to 51.2) |
| ≥1 month after Dose 3 | 1,580 | 6,211 | 2,029 | 5,762 | 42.0 (35.6 to 47.8) |
| Effectiveness against any symptomatic Omicron infection<sup>**</sup> |  |  |  |  |  |
| Dose 1 |  |  |  |  |  |
| 0-13 days after Dose 1 and no Dose 2 | 56 | 12,174 | 34 | 7,278 | 5.9 (-47.1 to 39.8) |
| ≥14 days after Dose 1 and no Dose 2 | 151 | 12,205 | 130 | 7,276 | 29.6 (9.6 to 45.2) |
| Dose 2 |  |  |  |  |  |
| 1-3 months after Dose 2 and no Dose 3 | 585 | 12,623 | 599 | 7,309 | 44.8 (37.2 to 51.5) |
| 4-6 months after Dose 2 and no Dose 3 | 2,479 | 12,590 | 1,605 | 7,333 | 12.1 (5.0 to 18.7) |
| ≥7 months after Dose 2 and no Dose 3 | 16,435 | 12,564 | 9,073 | 7,637 | -9.5 (-15.2 to -4.1) |
| Dose 3 (booster dose) |  |  |  |  |  |
| <1 month after Dose 3 | 1,326 | 12,777 | 1,300 | 7,275 | 49.9 (44.5 to 54.8) |
| ≥1 month after Dose 3 | 3,120 | 13,040 | 2,484 | 7,360 | 42.9 (37.7 to 47.6) |

Abbreviations: CI, confidence interval; PCR, polymerase chain reaction.
\*A symptomatic infection was defined as a PCR-positive nasopharyngeal swab conducted because of clinical suspicion due to presence of symptoms compatible with a respiratory tract infection.
<sup>†</sup>In each analysis for a specific time-since-vaccination stratum, we included only those vaccinated in this specific time-since-vaccination stratum and those unvaccinated. Only matched pairs of PCR-positive and PCR-negative persons, in which both members of the pair were either unvaccinated or fell within each time-since-vaccination stratum have been included in the corresponding vaccine effectiveness estimate. Thus, the number of cases (and controls) varied across time-since-vaccination analyses.
<sup>‡</sup>Vaccine effectiveness was estimated using the test-negative, case-control study design<sup>1,2</sup>.
<sup>§</sup>Cases and controls were matched one-to-two by sex, 10-year age group, nationality, and calendar week of PCR test.
<sup>¶</sup>Cases and controls were matched one-to-one by sex, 10-year age group, nationality, and calendar week of PCR test.
<sup>\*\*</sup>Cases and controls were matched two-to-one by sex, 10-year age group, nationality, and calendar week of PCR test.

**Supplementary Table 2.** Sensitivity analysis for the effectiveness of the mRNA-1273 vaccine against symptomatic SARS-CoV-2 BA.1 Omicron infection, BA.2 Omicron infection, and any Omicron infection*, adjusting for documented prior infection and health worker status in the conditional logistic regression.

| Sub-studies<sup>†</sup> | Cases<br>(PCR-positive) |  | Controls<br>(PCR-negative) |  | Effectiveness in %<br>(95% CI)<sup>‡</sup> |
| --- | --- | --- | --- | --- | --- |
|  | Vaccinated | Unvaccinated | Vaccinated | Unvaccinated |  |
| Effectiveness against symptomatic BA.1 Omicron infection<sup>§</sup> |  |  |  |  |  |
| Dose 1 |  |  |  |  |  |
| 0-13 days after Dose 1 and no Dose 2 | 3 | 1,942 | 8 | 3,400 | 45.8 (-115.2 to 86.3) |
| ≥14 days after Dose 1 and no Dose 2 | 14 | 1,942 | 19 | 3,405 | -40.1 (-191.7 to 32.7) |
| Dose 2 |  |  |  |  |  |
| 1-3 months after Dose 2 and no Dose 3 | 6 | 1,943 | 27 | 3,396 | 71.0 (22.6 to 89.1) |
| 4-6 months after Dose 2 and no Dose 3 | 289 | 1,976 | 667 | 3,377 | 27.1 (13.6 to 38.5) |
| ≥7 months after Dose 2 and no Dose 3 | 1,125 | 1,999 | 1,847 | 3,638 | -16.8 (-31.1 to -4.1) |
| Dose 3 (booster dose) |  |  |  |  |  |
| <1 month after Dose 3 | 55 | 1,951 | 182 | 3,377 | 48.9 (27.9 to 63.8) |
| ≥1 month after Dose 3 | 36 | 1,953 | 102 | 3,396 | 46.0 (17.2 to 64.8) |
| Effectiveness against symptomatic BA.2 Omicron infection<sup>¶</sup> |  |  |  |  |  |
| Dose 1 |  |  |  |  |  |
| 0-13 days after Dose 1 and no Dose 2 | 8 | 5,651 | 10 | 5,649 | 3.0 (-151.5 to 62.6) |
| ≥14 days after Dose 1 and no Dose 2 | 31 | 5,645 | 27 | 5,649 | -19.9 (-104.9 to 29.9) |
| Dose 2 |  |  |  |  |  |
| 1-3 months after Dose 2 and no Dose 3 | 26 | 5,664 | 40 | 5,650 | 39.7 (-1.2 to 64.1) |
| 4-6 months after Dose 2 and no Dose 3 | 989 | 5,756 | 1,059 | 5,686 | 6.0 (-4.9 to 15.8) |
| ≥7 months after Dose 2 and no Dose 3 | 2,917 | 5,627 | 2,686 | 5,858 | -24.0 (-34.5 to -14.3) |
| Dose 3 (booster dose) |  |  |  |  |  |
| <1 month after Dose 3 | 164 | 5,727 | 250 | 5,641 | 36.7 (20.9 to 49.2) |
| ≥1 month after Dose 3 | 92 | 5,709 | 149 | 5,652 | 42.5 (23.6 to 56.7) |
| Effectiveness against any symptomatic Omicron infection<sup>**</sup> |  |  |  |  |  |
| Dose 1 |  |  |  |  |  |
| 0-13 days after Dose 1 | 17 | 11,987 | 11 | 7,153 | 3.7 (-108.9 to 55.6) |
| ≥14 days after Dose 1 and no Dose 2 | 52 | 11,984 | 36 | 7,150 | 5.8 (-47.0 to 39.7) |
| Dose 2 |  |  |  |  |  |
| 1-3 months after Dose 2 and no Dose 3 | 47 | 12,014 | 51 | 7,151 | 39.7 (8.8 to 60.2) |
| 4-6 months after Dose 2 and no Dose 3 | 1,863 | 12,321 | 1,294 | 7,205 | 15.0 (7.0 to 22.3) |
| ≥7 months after Dose 2 and no Dose 3 | 5,820 | 12,144 | 3,112 | 7,374 | -17.0 (-25.0 to -9.5) |
| Dose 3 (booster dose) |  |  |  |  |  |
| <1 month after Dose 3 | 323 | 12,156 | 321 | 7,148 | 40.8 (29.6 to 50.3) |
| ≥1 month after Dose 3 | 169 | 12,100 | 181 | 7,155 | 48.3 (34.7 to 59.1) |

Abbreviations: CI, confidence interval; PCR, polymerase chain reaction.
\*A symptomatic infection was defined as a PCR-positive nasopharyngeal swab conducted because of clinical suspicion due to presence of symptoms compatible with a respiratory tract infection.
†In each analysis for a specific time-since-vaccination stratum, we included only those vaccinated in this specific time-since-vaccination stratum and those unvaccinated. Only matched pairs of PCR-positive and PCR-negative persons, in which both members of the pair were either unvaccinated or fell within each time-since-vaccination stratum have been included in the corresponding vaccine effectiveness estimate. Thus, the number of cases (and controls) varied across time-since-vaccination analyses.
‡Vaccine effectiveness was estimated using the test-negative, case-control study design<sup>1,2</sup>.
§Cases and controls were matched one-to-two by sex, 10-year age group, nationality, and calendar week of PCR test.
¶Cases and controls were matched one-to-one by sex, 10-year age group, nationality, and calendar week of PCR test.
\*\*Cases and controls were matched two-to-one by sex, 10-year age group, nationality, and calendar week of PCR test.

**Supplementary Table 3.** Sensitivity analysis for the effectiveness of the BNT162b2 vaccine against symptomatic SARS-CoV-2 BA.1 Omicron infection, BA.2 Omicron infection, and any Omicron infection*, after excluding children <12 years of age.

| Sub-studies<sup>†</sup> | Cases<br>(PCR-positive) |  | Controls<br>(PCR-negative) |  | Effectiveness in %<br>(95% CI)<sup>‡</sup> |
| --- | --- | --- | --- | --- | --- |
|  | Vaccinated | Unvaccinated | Vaccinated | Unvaccinated |  |
| Effectiveness against symptomatic BA.1 Omicron infection<sup>§</sup> |  |  |  |  |  |
| Dose 1 |  |  |  |  |  |
| 0-13 days after Dose 1 and no Dose 2 | 10 | 1,155 | 21 | 1,886 | 23.4 (-64.2 to 64.2) |
| ≥14 days after Dose 1 and no Dose 2 | 25 | 1,155 | 65 | 1,874 | 37.6 (0.3 to 60.9) |
| Dose 2 |  |  |  |  |  |
| 1-3 months after Dose 2 and no Dose 3 | 127 | 1,181 | 351 | 1,858 | 45.1 (31.1 to 56.3) |
| 4-6 months after Dose 2 and no Dose 3 | 499 | 1,195 | 960 | 1,921 | 14.1 (1.3 to 25.2) |
| ≥7 months after Dose 2 and no Dose 3 | 3,564 | 1,251 | 6,001 | 2,380 | -17.3 (-28.0 to -7.5) |
| Dose 3 (booster dose) |  |  |  |  |  |
| <1 month after Dose 3 | 181 | 1,192 | 637 | 1,754 | 61.0 (52.7 to 67.9) |
| ≥1 month after Dose 3 | 483 | 1,218 | 1,171 | 1,847 | 42.4 (33.2 to 50.4) |
| Effectiveness against symptomatic BA.2 Omicron infection<sup>¶</sup> |  |  |  |  |  |
| Dose 1 |  |  |  |  |  |
| 0-13 days after Dose 1 and no Dose 2 | 26 | 2,973 | 30 | 2,969 | 13.3 (-46.5 to 48.7) |
| ≥14 days after Dose 1 and no Dose 2 | 67 | 2,996 | 97 | 2,966 | 31.6 (6.2 to 50.1) |
| Dose 2 |  |  |  |  |  |
| 1-3 months after Dose 2 and no Dose 3 | 257 | 3,199 | 488 | 2,968 | 51.4 (42.9 to 58.7) |
| 4-6 months after Dose 2 and no Dose 3 | 1,233 | 3,121 | 1,319 | 3,035 | 10.0 (0.8 to 18.3) |
| ≥7 months after Dose 2 and no Dose 3 | 8,001 | 3,064 | 7,742 | 3,323 | -13.7 (-21.0 to -6.9) |
| Dose 3 (booster dose) |  |  |  |  |  |
| <1 month after Dose 3 | 705 | 3,265 | 1,057 | 2,913 | 44.5 (37.6 to 50.6) |
| ≥1 month after Dose 3 | 1,604 | 3,412 | 2,025 | 2,991 | 38.9 (32.7 to 44.5) |
| Effectiveness against any symptomatic Omicron infection<sup>**</sup> |  |  |  |  |  |
| Dose 1 |  |  |  |  |  |
| 0-13 days after Dose 1 and no Dose 2 | 55 | 6,043 | 32 | 3,619 | 3.4 (-51.0 to 38.2) |
| ≥14 days after Dose 1 and no Dose 2 | 133 | 6,097 | 128 | 3,616 | 37.2 (19.4 to 51.1) |
| Dose 2 |  |  |  |  |  |
| 1-3 months after Dose 2 and no Dose 3 | 616 | 6,466 | 588 | 3,656 | 43.8 (36.3 to 50.5) |
| 4-6 months after Dose 2 and no Dose 3 | 2,511 | 6,413 | 1,586 | 3,689 | 13.1 (6.0 to 19.7) |
| ≥7 months after Dose 2 and no Dose 3 | 16,552 | 6,302 | 9,025 | 4,014 | -15.0 (-21.0 to -9.4) |
| Dose 3 (booster dose) |  |  |  |  |  |
| <1 month after Dose 3 | 1,323 | 6,648 | 1,278 | 3,634 | 48.0 (42.7 to 52.8) |
| ≥1 month after Dose 3 | 3,113 | 6,919 | 2,494 | 3,689 | 40.7 (35.8 to 45.3) |

Abbreviations: CI, confidence interval; PCR, polymerase chain reaction.
\*A symptomatic infection was defined as a PCR-positive nasopharyngeal swab conducted because of clinical suspicion due to presence of symptoms compatible with a respiratory tract infection.
<sup>†</sup>In each analysis for a specific time-since-vaccination stratum, we included only those vaccinated in this specific time-since-vaccination stratum and those unvaccinated. Only matched pairs of PCR-positive and PCR-negative persons, in which both members of the pair were either unvaccinated or fell within each time-since-vaccination stratum have been included in the corresponding vaccine effectiveness estimate. Thus, the number of cases (and controls) varied across time-since-vaccination analyses.
<sup>‡</sup>Vaccine effectiveness was estimated using the test-negative, case-control study design<sup>1,2</sup>.
<sup>§</sup>Cases and controls were matched one-to-two by sex, 10-year age group, nationality, and calendar week of PCR test.
<sup>¶</sup>Cases and controls were matched one-to-one by sex, 10-year age group, nationality, and calendar week of PCR test.
\*\*Cases and controls were matched two-to-one by sex, 10-year age group, nationality, and calendar week of PCR test.

**Supplementary Table 4.** Sensitivity analysis for the effectiveness of the mRNA-1273 vaccine against symptomatic SARS-CoV-2 BA.1 Omicron infection, BA.2 Omicron infection, and any Omicron infection*, after excluding children <12 years of age.

| Sub-studies<sup>†</sup> | Cases<br>(PCR-positive) |  | Controls<br>(PCR-negative) |  | Effectiveness in %<br>(95% CI)<sup>‡</sup> |
| --- | --- | --- | --- | --- | --- |
|  | Vaccinated | Unvaccinated | Vaccinated | Unvaccinated |  |
| Effectiveness against symptomatic BA.1 Omicron infection<sup>§</sup> |  |  |  |  |  |
| Dose 1 |  |  |  |  |  |
| 0-13 days after Dose 1 and no Dose 2 | 3 | 1,128 | 9 | 1,830 | 48.5 (-93.2 to 86.2) |
| ≥14 days after Dose 1 and no Dose 2 | 14 | 1,128 | 17 | 1,838 | -29.5 (-168.7 to 37.6) |
| Dose 2 |  |  |  |  |  |
| 1-3 months after Dose 2 and no Dose 3 | 6 | 1,129 | 30 | 1,824 | 70.1 (27.2 to 87.7) |
| 4-6 months after Dose 2 and no Dose 3 | 283 | 1,169 | 664 | 1,807 | 33.7 (21.6 to 44.0) |
| ≥7 months after Dose 2 and no Dose 3 | 1,125 | 1,184 | 1,830 | 2,074 | -10.6 (-23.6 to 1.0) |
| Dose 3 (booster dose) |  |  |  |  |  |
| <1 month after Dose 3 | 54 | 1,139 | 184 | 1,807 | 52.0 (33.4 to 65.4) |
| ≥1 month after Dose 3 | 36 | 1,139 | 101 | 1,827 | 43.4 (15.5 to 62.1) |
| Effectiveness against symptomatic BA.2 Omicron infection<sup>§</sup> |  |  |  |  |  |
| Dose 1 |  |  |  |  |  |
| 0-13 days after Dose 1 and no Dose 2 | 8 | 2,876 | 9 | 2,875 | 11.1 (-130.3 to 65.7) |
| ≥14 days after Dose 1 and no Dose 2 | 22 | 2,879 | 29 | 2,872 | 25.0 (-32.1 to 57.4) |
| Dose 2 |  |  |  |  |  |
| 1-3 months after Dose 2 and no Dose 3 | 18 | 2,897 | 37 | 2,878 | 51.4 (14.6 to 72.3) |
| 4-6 months after Dose 2 and no Dose 3 | 945 | 3,022 | 1,059 | 2,908 | 16.1 (6.4 to 24.7) |
| ≥7 months after Dose 2 and no Dose 3 | 2,861 | 2,906 | 2,676 | 3,091 | -16.0 (-25.6 to -7.3) |
| Dose 3 (booster dose) |  |  |  |  |  |
| <1 month after Dose 3 | 169 | 2,947 | 265 | 2,851 | 40.7 (26.9 to 51.9) |
| ≥1 month after Dose 3 | 84 | 2,941 | 153 | 2,872 | 47.9 (31.1 to 60.6) |
| Effectiveness against any symptomatic Omicron infection<sup>**</sup> |  |  |  |  |  |
| Dose 1 |  |  |  |  |  |
| 0-13 days after Dose 1 and no Dose 2 | 19 | 5,856 | 9 | 3,495 | -17.6 (-161.4 to 47.1) |
| ≥14 days after Dose 1 and no Dose 2 | 40 | 5,868 | 36 | 3,490 | 28.9 (-12.3 to 54.9) |
| Dose 2 |  |  |  |  |  |
| 1-3 months after Dose 2 and no Dose 3 | 43 | 5,890 | 46 | 3,497 | 45.2 (16.0 to 64.2) |
| 4-6 months after Dose 2 and no Dose 3 | 1,878 | 6,175 | 1,293 | 3,544 | 18.2 (10.6 to 25.1) |
| ≥7 months after Dose 2 and no Dose 3 | 5,832 | 6,001 | 3,107 | 3,716 | -15.0 (-22.7 to -7.8) |
| Dose 3 (booster dose) |  |  |  |  |  |
| <1 month after Dose 3 | 327 | 6,027 | 320 | 3,490 | 44.4 (33.8 to 53.2) |
| ≥1 month after Dose 3 | 174 | 5,965 | 179 | 3,498 | 46.0 (32.4 to 56.9) |

Abbreviations: CI, confidence interval; PCR, polymerase chain reaction.
\*A symptomatic infection was defined as a PCR-positive nasopharyngeal swab conducted because of clinical suspicion due to presence of symptoms compatible with a respiratory tract infection.
<sup>‡</sup>In each analysis for a specific time-since-vaccination stratum, we included only those vaccinated in this specific time-since-vaccination stratum and those unvaccinated. Only matched pairs of PCR-positive and PCR-negative persons, in which both members of the pair were either unvaccinated or fell within each time-since-vaccination stratum have been included in the corresponding vaccine effectiveness estimate. Thus, the number of cases (and controls) varied across time-since-vaccination analyses.
<sup>§</sup>Vaccine effectiveness was estimated using the test-negative, case-control study design<sup>1,2</sup>.
<sup>§</sup>Cases and controls were matched one-to-two by sex, 10-year age group, nationality, and calendar week of PCR test.
<sup>§</sup>Cases and controls were matched one-to-one by sex, 10-year age group, nationality, and calendar week of PCR test.
<sup>\*\*</sup>Cases and controls were matched two-to-one by sex, 10-year age group, nationality, and calendar week of PCR test.

**Supplementary Table 5.** Sensitivity analysis for the effectiveness of the BNT162b2 vaccine against symptomatic SARS-CoV-2 BA.1 Omicron infection, BA.2 Omicron infection, and any Omicron infection*, after excluding individuals <20 years of age.

| Sub-studies<sup>†</sup> | Cases<br>(PCR-positive) |  | Controls<br>(PCR-negative) |  | Effectiveness in %<br>(95% CI)<sup>‡</sup> |
| --- | --- | --- | --- | --- | --- |
|  | Vaccinated | Unvaccinated | Vaccinated | Unvaccinated |  |
| Effectiveness against symptomatic BA.1 Omicron infection<sup>§</sup> |  |  |  |  |  |
| Dose 1 |  |  |  |  |  |
| 0-13 days after Dose 1 and no Dose 2 | 9 | 1,034 | 19 | 1,677 | 24.9 (-71.7 to 67.2) |
| ≥14 days after Dose 1 and no Dose 2 | 18 | 1,035 | 56 | 1,669 | 47.6 (10.0 to 69.5) |
| Dose 2 |  |  |  |  |  |
| 1-3 months after Dose 2 and no Dose 3 | 87 | 1,050 | 259 | 1,648 | 48.1 (32.4 to 60.1) |
| 4-6 months after Dose 2 and no Dose 3 | 348 | 1,055 | 628 | 1,710 | 9.4 (-6.1 to 22.7) |
| ≥7 months after Dose 2 and no Dose 3 | 3,203 | 1,115 | 5,357 | 2,130 | -18.4 (-29.9 to -7.9) |
| Dose 3 (booster dose) |  |  |  |  |  |
| <1 month after Dose 3 | 177 | 1,070 | 607 | 1,570 | 60.8 (52.0 to 68.0) |
| ≥1 month after Dose 3 | 480 | 1,096 | 1,141 | 1,661 | 40.0 (30.4 to 48.4) |
| Effectiveness against symptomatic BA.2 Omicron infection<sup>¶</sup> |  |  |  |  |  |
| Dose 1 |  |  |  |  |  |
| 0-13 days after Dose 1 and no Dose 2 | 26 | 2,674 | 26 | 2,674 | 0.0 (-72.2 to 41.9) |
| ≥14 days after Dose 1 and no Dose 2 | 58 | 2,699 | 88 | 2,669 | 35.7 (9.5 to 54.3) |
| Dose 2 |  |  |  |  |  |
| 1-3 months after Dose 2 and no Dose 3 | 196 | 2,826 | 352 | 2,670 | 48.0 (37.4 to 56.8) |
| 4-6 months after Dose 2 and no Dose 3 | 955 | 2,701 | 908 | 2,748 | -7.5 (-19.9 to 3.6) |
| ≥7 months after Dose 2 and no Dose 3 | 7,404 | 2,605 | 6,986 | 3,023 | -27.0 (-35.7 to -18.8) |
| Dose 3 (booster dose) |  |  |  |  |  |
| <1 month after Dose 3 | 724 | 2,937 | 1,028 | 2,633 | 41.5 (34.1 to 48.1) |
| ≥1 month after Dose 3 | 1,612 | 3,096 | 2,003 | 2,705 | 36.6 (30.2 to 42.4) |
| Effectiveness against any symptomatic Omicron infection<sup>**</sup> |  |  |  |  |  |
| Dose 1 |  |  |  |  |  |
| 0-13 days after Dose 1 and no Dose 2 | 42 | 5,434 | 27 | 3,261 | 12.9 (-42.3 to 46.7) |
| ≥14 days after Dose 1 and no Dose 2 | 107 | 5,484 | 107 | 3,261 | 38.7 (19.3 to 53.5) |
| Dose 2 |  |  |  |  |  |
| 1-3 months after Dose 2 and no Dose 3 | 458 | 5,713 | 416 | 3,291 | 39.5 (30.0 to 47.7) |
| 4-6 months after Dose 2 and no Dose 3 | 1,860 | 5,561 | 1,065 | 3,317 | -0.04 (-9.5 to 8.6) |
| ≥7 months after Dose 2 and no Dose 3 | 15,089 | 5,543 | 8,167 | 3,610 | -19.6 (-26.2 to -13.3) |
| Dose 3 (booster dose) |  |  |  |  |  |
| <1 month after Dose 3 | 1,316 | 6,012 | 1,270 | 3,266 | 47.4 (42.0 to 52.3) |
| ≥1 month after Dose 3 | 3,107 | 6,283 | 2,458 | 3,349 | 38.9 (33.9 to 43.6) |

Abbreviations: CI, confidence interval; PCR, polymerase chain reaction.
\*A symptomatic infection was defined as a PCR-positive nasopharyngeal swab conducted because of clinical suspicion due to presence of symptoms compatible with a respiratory tract infection.
<sup>‡</sup>In each analysis for a specific time-since-vaccination stratum, we included only those vaccinated in this specific time-since-vaccination stratum and those unvaccinated. Only matched pairs of PCR-positive and PCR-negative persons, in which both members of the pair were either unvaccinated or fell within each time-since-vaccination stratum have been included in the corresponding vaccine effectiveness estimate. Thus, the number of cases (and controls) varied across time-since-vaccination analyses.
<sup>§</sup>Vaccine effectiveness was estimated using the test-negative, case-control study design<sup>1,2</sup>.
<sup>¶</sup>Cases and controls were matched one-to-two by sex, 10-year age group, nationality, and calendar week of PCR test.
<sup>¶</sup>Cases and controls were matched one-to-one by sex, 10-year age group, nationality, and calendar week of PCR test.
\*\*Cases and controls were matched two-to-one by sex, 10-year age group, nationality, and calendar week of PCR test.

**Supplementary Table 6.** Sensitivity analysis for the effectiveness of the mRNA-1273 vaccine against symptomatic SARS-CoV-2 BA.1 Omicron infection, BA.2 Omicron infection, and any Omicron infection*, after excluding individuals <20 years of age.

| Sub-studies<sup>†</sup> | Cases<br>(PCR-positive) |  | Controls<br>(PCR-negative) |  | Effectiveness in %<br>(95% CI)<sup>‡</sup> |
| --- | --- | --- | --- | --- | --- |
|  | Vaccinated | Unvaccinated | Vaccinated | Unvaccinated |  |
| Effectiveness against symptomatic BA.1 Omicron infection<sup>§</sup> |  |  |  |  |  |
| Dose 1 |  |  |  |  |  |
| 0-13 days after Dose 1 and no Dose 2 | 3 | 1,011 | 7 | 1,630 | 40.8 (-132.7 to 85.0) |
| ≥14 days after Dose 1 and no Dose 2 | 14 | 1,011 | 17 | 1,637 | -28.3 (-162.9 to 37.4) |
| Dose 2 |  |  |  |  |  |
| 1-3 months after Dose 2 and no Dose 3 | 6 | 1,012 | 27 | 1,625 | 65.1 (14.9 to 85.7) |
| 4-6 months after Dose 2 and no Dose 3 | 282 | 1,043 | 673 | 1,580 | 34.6 (22.8 to 44.6) |
| ≥7 months after Dose 2 and no Dose 3 | 1,106 | 1,070 | 1,807 | 1,873 | -9.7 (-22.8 to 1.9) |
| Dose 3 (booster dose) |  |  |  |  |  |
| <1 month after Dose 3 | 54 | 1,021 | 178 | 1,610 | 50.6 (31.1 to 64.6) |
| ≥1 month after Dose 3 | 37 | 1,021 | 91 | 1,636 | 32.5 (-0.4 to 54.6) |
| Effectiveness against symptomatic BA.2 Omicron infection<sup>¶</sup> |  |  |  |  |  |
| Dose 1 |  |  |  |  |  |
| 0-13 days after Dose 1 and no Dose 2 | 8 | 2,593 | 7 | 2,594 | -14.3 (-215.2 to 58.6) |
| ≥14 days after Dose 1 and no Dose 2 | 28 | 2,591 | 27 | 2,592 | -3.8 (-77.9 to 39.4) |
| Dose 2 |  |  |  |  |  |
| 1-3 months after Dose 2 and no Dose 3 | 22 | 2,609 | 42 | 2,589 | 48.8 (13.3 to 69.7) |
| 4-6 months after Dose 2 and no Dose 3 | 950 | 2,719 | 1,025 | 2,644 | 10.9 (0.7 to 20.1) |
| ≥7 months after Dose 2 and no Dose 3 | 2,866 | 2,587 | 2,624 | 2,829 | -21.9 (-32.0 to -12.6) |
| Dose 3 (booster dose) |  |  |  |  |  |
| <1 month after Dose 3 | 167 | 2,665 | 255 | 2,577 | 37.9 (23.6 to 49.6) |
| ≥1 month after Dose 3 | 93 | 2,648 | 150 | 2,591 | 41.6 (23.1 to 55.7) |
| Effectiveness against any symptomatic Omicron infection<sup>**</sup> |  |  |  |  |  |
| Dose 1 |  |  |  |  |  |
| 0-13 days after Dose 1 and no Dose 2 | 17 | 5,266 | 8 | 3,151 | -16.3 (-171.0 to 50.1) |
| ≥14 days after Dose 1 and no Dose 2 | 47 | 5,271 | 31 | 3,151 | 7.2 (-47.3 to 41.5) |
| Dose 2 |  |  |  |  |  |
| 1-3 months after Dose 2 and no Dose 3 | 46 | 5,294 | 47 | 3,150 | 42.8 (13.0 to 62.5) |
| 4-6 months after Dose 2 and no Dose 3 | 1,877 | 5,551 | 1,270 | 3,203 | 16.0 (8.2 to 23.2) |
| ≥7 months after Dose 2 and no Dose 3 | 5,763 | 5,402 | 3,076 | 3,357 | -14.4 (-22.1 to -7.2) |
| Dose 3 (booster dose) |  |  |  |  |  |
| <1 month after Dose 3 | 296 | 5,461 | 319 | 3,142 | 47.5 (37.7 to 55.8) |
| ≥1 month after Dose 3 | 166 | 5,376 | 170 | 3,159 | 44.4 (30.2 to 55.8) |

Abbreviations: CI, confidence interval; PCR, polymerase chain reaction.
<sup>‡</sup>A symptomatic infection was defined as a PCR-positive nasopharyngeal swab conducted because of clinical suspicion due to presence of symptoms compatible with a respiratory tract infection.
<sup>§</sup>In each analysis for a specific time-since-vaccination stratum, we included only those vaccinated in this specific time-since-vaccination stratum and those unvaccinated. Only matched pairs of PCR-positive and PCR-negative persons, in which both members of the pair were either unvaccinated or fell within each time-since-vaccination stratum have been included in the corresponding vaccine effectiveness estimate. Thus, the number of cases (and controls) varied across time-since-vaccination analyses.
<sup>¶</sup>Vaccine effectiveness was estimated using the test-negative, case-control study design<sup>1,2</sup>.
<sup>§</sup>Cases and controls were matched one-to-two by sex, 10-year age group, nationality, and calendar week of PCR test.
<sup>¶</sup>Cases and controls were matched one-to-one by sex, 10-year age group, nationality, and calendar week of PCR test.
<sup>\*\*</sup>Cases and controls were matched two-to-one by sex, 10-year age group, nationality, and calendar week of PCR test.

**Supplementary Table 7.**
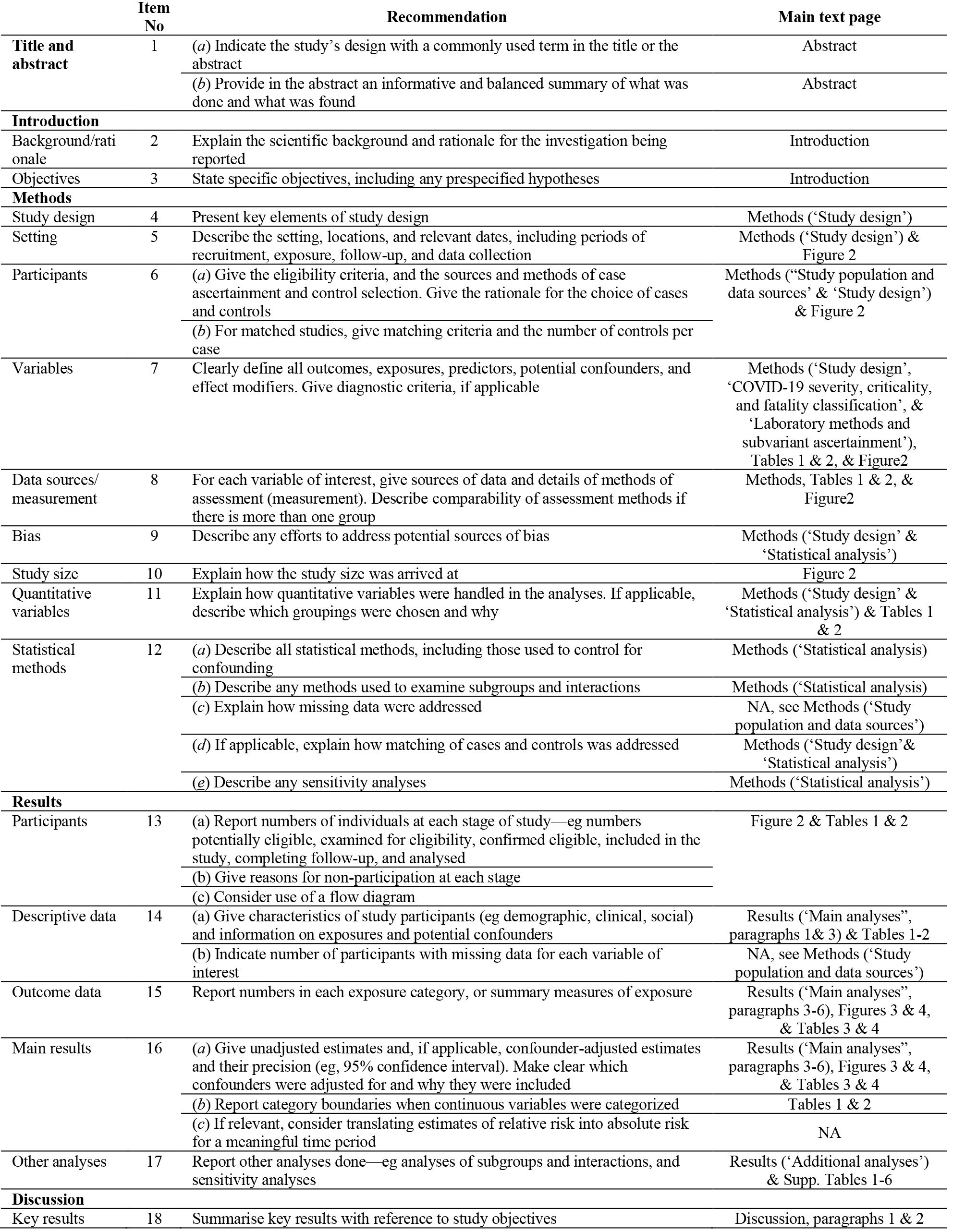
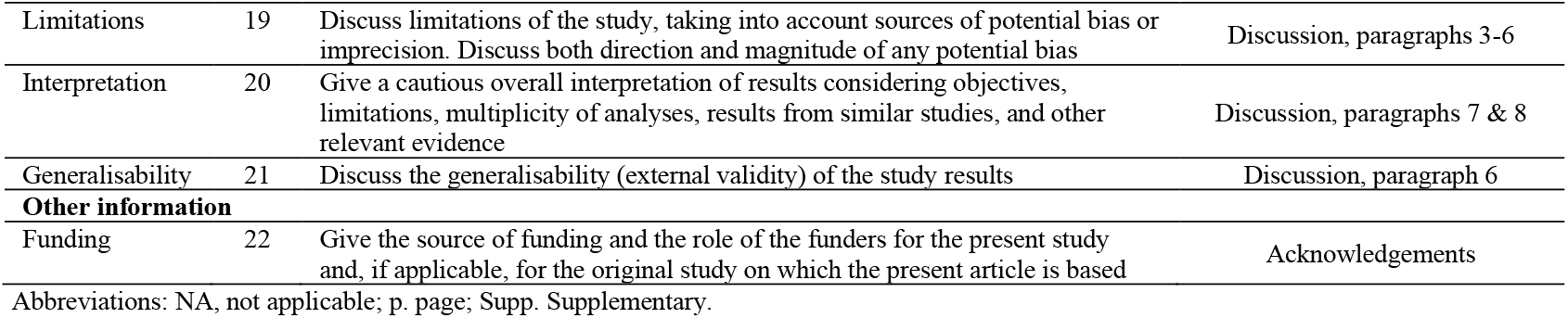
STROBE checklist for case-control studies.

|  | Item No | Recommendation | Main text page |
| --- | --- | --- | --- |
| Title and abstract | 1 | (a) Indicate the study’s design with a commonly used term in the title or the abstract | Abstract |
|  |  | (b) Provide in the abstract an informative and balanced summary of what was done and what was found | Abstract |
| Introduction |  |  |  |
| Background/rationale | 2 | Explain the scientific background and rationale for the investigation being reported | Introduction |
| Objectives | 3 | State specific objectives, including any prespecified hypotheses | Introduction |
| Methods |  |  |  |
| Study design | 4 | Present key elements of study design | Methods (‘Study design’) |
| Setting | 5 | Describe the setting, locations, and relevant dates, including periods of recruitment, exposure, follow-up, and data collection | Methods (‘Study design’) & Figure 2 |
| Participants | 6 | (a) Give the eligibility criteria, and the sources and methods of case ascertainment and control selection. Give the rationale for the choice of cases and controls<br>(b) For matched studies, give matching criteria and the number of controls per case | Methods (‘Study population and data sources’ & ‘Study design’) & Figure 2 |
| Variables | 7 | Clearly define all outcomes, exposures, predictors, potential confounders, and effect modifiers. Give diagnostic criteria, if applicable | Methods (‘Study design’, ‘COVID-19 severity, criticality, and fatality classification’, & ‘Laboratory methods and subvariant ascertainment’), Tables 1 & 2, & Figure2 |
| Data sources/measurement | 8 | For each variable of interest, give sources of data and details of methods of assessment (measurement). Describe comparability of assessment methods if there is more than one group | Methods, Tables 1 & 2, & Figure2 |
| Bias | 9 | Describe any efforts to address potential sources of bias | Methods (‘Study design’ & ‘Statistical analysis’) |
| Study size | 10 | Explain how the study size was arrived at | Figure 2 |
| Quantitative variables | 11 | Explain how quantitative variables were handled in the analyses. If applicable, describe which groupings were chosen and why | Methods (‘Study design’ & ‘Statistical analysis’) & Tables 1 & 2 |
| Statistical methods | 12 | (a) Describe all statistical methods, including those used to control for confounding | Methods (‘Statistical analysis’) |
|  |  | (b) Describe any methods used to examine subgroups and interactions | Methods (‘Statistical analysis’) |
|  |  | (c) Explain how missing data were addressed | NA, see Methods (‘Study population and data sources’) |
|  |  | (d) If applicable, explain how matching of cases and controls was addressed | Methods (‘Study design’& ‘Statistical analysis’) |
|  |  | (e) Describe any sensitivity analyses | Methods (‘Statistical analysis’) |
| Results |  |  |  |
| Participants | 13 | (a) Report numbers of individuals at each stage of study—eg numbers potentially eligible, examined for eligibility, confirmed eligible, included in the study, completing follow-up, and analysed<br>(b) Give reasons for non-participation at each stage<br>(c) Consider use of a flow diagram | Figure 2 & Tables 1 & 2 |
| Descriptive data | 14 | (a) Give characteristics of study participants (eg demographic, clinical, social) and information on exposures and potential confounders<br>(b) Indicate number of participants with missing data for each variable of interest | Results (‘Main analyses’, paragraphs 1& 3) & Tables 1-2<br>NA, see Methods (‘Study population and data sources’) |
| Outcome data | 15 | Report numbers in each exposure category, or summary measures of exposure | Results (‘Main analyses’, paragraphs 3-6), Figures 3 & 4, & Tables 3 & 4 |
| Main results | 16 | (a) Give unadjusted estimates and, if applicable, confounder-adjusted estimates and their precision (eg, 95% confidence interval). Make clear which confounders were adjusted for and why they were included | Results (‘Main analyses’, paragraphs 3-6), Figures 3 & 4, & Tables 3 & 4 |
|  |  | (b) Report category boundaries when continuous variables were categorized | Tables 1 & 2 |
|  |  | (c) If relevant, consider translating estimates of relative risk into absolute risk for a meaningful time period | NA |
| Other analyses | 17 | Report other analyses done—eg analyses of subgroups and interactions, and sensitivity analyses | Results (‘Additional analyses’) & Supp. Tables 1-6 |
| Discussion |  |  |  |
| Key results | 18 | Summarise key results with reference to study objectives | Discussion, paragraphs 1 & 2 |
| Limitations | 19 | Discuss limitations of the study, taking into account sources of potential bias or imprecision. Discuss both direction and magnitude of any potential bias | Discussion, paragraphs 3-6 |
| Interpretation | 20 | Give a cautious overall interpretation of results considering objectives, limitations, multiplicity of analyses, results from similar studies, and other relevant evidence | Discussion, paragraphs 7 & 8 |
| Generalisability | 21 | Discuss the generalisability (external validity) of the study results | Discussion, paragraph 6 |
| **Other information** |  |  |  |
| Funding | 22 | Give the source of funding and the role of the funders for the present study and, if applicable, for the original study on which the present article is based | Acknowledgements |

Abbreviations: NA, not applicable; p. page; Supp. Supplementary.

## References

1. World Health Organization. Tracking SARS-CoV-2 variants. Available from: https://www.who.int/en/activities/tracking-SARS-CoV-2-variants/. (2021).

2. National Project of Surveillance for Variants of Concern and Viral Genome Sequencing. Qatar viral genome sequencing data. Data on randomly collected samples. https://www.gisaid.org/phylodynamics/global/nextstrain/. (2021).

3. Altarawneh, H.N., et al. Protection against the Omicron Variant from Previous SARS-CoV-2 Infection. N Engl J Med (2022).

4. Chemaitelly, H., et al. Protection of Omicron sub-lineage infection against reinfection with another Omicron sub-lineage. medRxiv, 2022.2002.2024.22271440 (2022).

5. Abu-Raddad, L.J., et al. Effect of mRNA Vaccine Boosters against SARS-CoV-2 Omicron Infection in Qatar. New England Journal of Medicine (2022).

6. Kumar, S., Karuppanan, K. & Subramaniam, G. Omicron (BA.1) and Sub-Variants (BA.1, BA.2 and BA.3) of SARS-CoV-2 Spike Infectivity and Pathogenicity: A Comparative Sequence and Structural-based Computational Assessment. bioRxiv, 2022.2002.2011.480029 (2022).

7. Polack, F.P., et al. Safety and Efficacy of the BNT162b2 mRNA Covid-19 Vaccine. N Engl J Med 383, 2603–2615 (2020).

8. Baden, L.R., et al. Efficacy and Safety of the mRNA-1273 SARS-CoV-2 Vaccine. N Engl J Med 384, 403–416 (2021).

9. World Health Organization. COVID-19 clinical management: living guidance. Available from: https://www.who.int/publications/i/item/WHO-2019-nCoV-clinical-2021-1. Accessed on: May 31, 2021. (2021).

10. World Health Organization. International guidelines for certification and classification (coding) of COVID-19 as cause of death. Available from: https://www.who.int/classifications/icd/Guidelines_Cause_of_Death_COVID-19-20200420-EN.pdf?ua=1. Document Number: WHO/HQ/DDI/DNA/CAT. Accessed on May 31, 2021. (2021).

11. Jackson, M.L. & Nelson, J.C. The test-negative design for estimating influenza vaccine effectiveness. Vaccine 31, 2165–2168 (2013).

12. Verani, J.R., et al. Case-control vaccine effectiveness studies: Preparation, design, and enrollment of cases and controls. Vaccine 35, 3295–3302 (2017).

13. Chemaitelly, H., et al. Waning of BNT162b2 Vaccine Protection against SARS-CoV-2 Infection in Qatar. N Engl J Med 385, e83 (2021).

14. Abu-Raddad, L.J., Chemaitelly, H., Bertollini, R. & National Study Group forCovid Vaccination. Waning mRNA-1273 Vaccine Effectiveness against SARS-CoV-2 Infection in Qatar. N Engl J Med (2022).

15. Abu-Raddad, L.J., et al. Characterizing the Qatar advanced-phase SARS-CoV-2 epidemic. Sci Rep 11, 6233 (2021).

16. Ayoub, H.H., et al. Mathematical modeling of the SARS-CoV-2 epidemic in Qatar and its impact on the national response to COVID-19. J Glob Health 11, 05005 (2021).

17. Coyle, P.V., et al. SARS-CoV-2 seroprevalence in the urban population of Qatar: An analysis of antibody testing on a sample of 112,941 individuals. iScience 24, 102646 (2021).

18. Al-Thani, M.H., et al. SARS-CoV-2 Infection Is at Herd Immunity in the Majority Segment of the Population of Qatar. Open Forum Infect Dis 8, ofab221 (2021).

19. Jeremijenko, A., et al. Herd Immunity against Severe Acute Respiratory Syndrome Coronavirus 2 Infection in 10 Communities, Qatar. Emerg Infect Dis 27, 1343–1352 (2021).

20. Wolter, N., et al. Early assessment of the clinical severity of the SARS-CoV-2 omicron variant in South Africa: a data linkage study. Lancet 399, 437–446 (2022).

21. Seedat, S., et al. SARS-CoV-2 infection hospitalization, severity, criticality, and fatality rates in Qatar. Sci Rep 11, 18182 (2021).

22. UK Health Security Agency. SARS-CoV-2 variants of concern and variants under investigation in England: Technical briefing 34. (England, 2022).

23. Abu-Raddad, L.J., et al. Introduction and expansion of the SARS-CoV-2 B.1.1.7 variant and reinfections in Qatar: A nationally representative cohort study. PLoS Med 18, e1003879 (2021).

24. Challen, R., et al. Risk of mortality in patients infected with SARS-CoV-2 variant of concern 202012/1: matched cohort study. BMJ 372, 579 (2021).

25. Makhoul M., et al. Epidemiological impact of SARS-CoV-2 vaccination: Mathematical modeling analyses. Vaccines 8(2020).

26. Usherwood, T., LaJoie, Z. & Srivastava, V. A model and predictions for COVID-19 considering population behavior and vaccination. Scientific Reports 11, 12051 (2021).

27. Andersson, O., Campos-Mercade, P., Meier, A.N. & Wengström, E. Anticipation of COVID-19 Vaccines Reduces Social Distancing. https://papers.ssrn.com/sol3/papers.cfm?abstract_id=3765329. SSRN (2021).

28. Ray, G.T., et al. Depletion-of-susceptibles Bias in Analyses of Intra-season Waning of Influenza Vaccine Effectiveness. Clin Infect Dis 70, 1484–1486 (2020).

29. Abu-Raddad, L.J., et al. Pfizer-BioNTech mRNA BNT162b2 Covid-19 vaccine protection against variants of concern after one versus two doses. J Travel Med 28(2021).

30. Chemaitelly, H., et al. mRNA-1273 COVID-19 vaccine effectiveness against the B.1.1.7 and B.1.351 variants and severe COVID-19 disease in Qatar. Nat Med 27, 1614–1621 (2021).

31. Abu-Raddad, L.J., Chemaitelly, H., Bertollini, R. & National Study Group for Covid Vaccination. Effectiveness of mRNA-1273 and BNT162b2 Vaccines in Qatar. N Engl J Med (2022).

32. Abu-Raddad, L.J., Chemaitelly, H., Butt, A.A. & National Study Group for Covid Vaccination. Effectiveness of the BNT162b2 Covid-19 Vaccine against the B.1.1.7 and B.1.351 Variants. N Engl J Med 385, 187–189 (2021).

33. Butt, A.A., et al. SARS-CoV-2 vaccine effectiveness in preventing confirmed infection in pregnant women. J Clin Invest 131(2021).

34. Lopez Bernal, J., et al. Effectiveness of Covid-19 Vaccines against the B.1.617.2 (Delta) Variant. N Engl J Med 385, 585–594 (2021).

35. Planning and Statistics Authority-State of Qatar. Qatar Monthly Statistics. Available from: https://www.psa.gov.qa/en/pages/default.aspx. Accessed on: May 26, 2020. (2020).

36. Andrews, N., et al. Covid-19 Vaccine Effectiveness against the Omicron (B.1.1.529) Variant. New England Journal of Medicine (2022).

37. Buchan, S.A., et al. Effectiveness of COVID-19 vaccines against Omicron or Delta infection. medRxiv, 2021.2012.2030.21268565 (2022).

38. Hansen, C.H., et al. Vaccine effectiveness against SARS-CoV-2 infection with the Omicron or Delta variants following a two-dose or booster BNT162b2 or mRNA-1273 vaccination series: A Danish cohort study. medRxiv, 2021.2012.2020.21267966 (2021).

39. Patalon, T., et al. Waning Effectiveness of the Third Dose of the BNT162b2 mRNA COVID-19 Vaccine. medRxiv, 2022.2002.2025.22271494 (2022).

40. Šmíd, M., et al. Protection by vaccines and previous infection against the Omicron variant of SARS-CoV-2. medRxiv, 2022.2002.2024.22271396 (2022).

41. Tseng, H.F., et al. Effectiveness of mRNA-1273 against SARS-CoV-2 omicron and delta variants. medRxiv, 2022.2001.2007.22268919 (2022).

42. Vogels, C., Fauver, J. & Grubaugh, N. Multiplexed RT-qPCR to screen for SARS-COV-2 B.1.1.7, B.1.351, and P.1 variants of concern V.3. dx.doi.org/10.17504/protocols.io.br9vm966. (2021).

43. Benslimane, F.M., et al. One Year of SARS-CoV-2: Genomic Characterization of COVID-19 Outbreak in Qatar. Front Cell Infect Microbiol 11, 768883 (2021).

44. Hasan, M.R., et al. Real-Time SARS-CoV-2 Genotyping by High-Throughput Multiplex PCR Reveals the Epidemiology of the Variants of Concern in Qatar. Int J Infect Dis 112, 52–54 (2021).

45. Saththasivam, J., et al. COVID-19 (SARS-CoV-2) outbreak monitoring using wastewater-based epidemiology in Qatar. Sci Total Environ 774, 145608 (2021).

46. Thermo Fisher Scientific. TaqPath™ COVID-19 CE-IVD RT-PCR Kit instructions for use. Available from: https://assets.thermofisher.com/TFS-Assets/LSG/manuals/MAN0019215_TaqPathCOVID-19_CE-IVD_RT-PCR%20Kit_IFU.pdf. Accessed on December 02, 2020. (2020).

47. Austin, P.C. Using the Standardized Difference to Compare the Prevalence of a Binary Variable Between Two Groups in Observational Research. Communications in Statistics-Simulation and Computation 38, 1228–1234 (2009).

48. Jacoby, P. & Kelly, H. Is it necessary to adjust for calendar time in a test negative design?: Responding to: Jackson ML, Nelson JC. The test negative design for estimating influenza vaccine effectiveness. Vaccine 2013;31(April (17)):2165-8. Vaccine 32, 2942 (2014).

49. Pearce, N. Analysis of matched case-control studies. BMJ 352, i969 (2016).

50. Rothman, K.J., Greenland, S. & Lash, T.L. Modern epidemiology, (Wolters Kluwer Health/Lippincott Williams & Wilkins, Philadelphia, 2008).

51. StataCorp. Stata Statistical Software: Release 17. College Station, TX: StataCorp LLC. (2021).

## References

1. Jackson, M.L. & Nelson, J.C. The test-negative design for estimating influenza vaccine effectiveness. Vaccine 31, 2165–2168 (2013).

2. Verani, J.R., et al. Case-control vaccine effectiveness studies: Preparation, design, and enrollment of cases and controls. Vaccine 35, 3295–3302 (2017).

